# Does the choice of stepping intensity metric influence dose-response associations with mortality? A UK population cohort study of 70,174 adults

**DOI:** 10.1101/2024.09.10.24313453

**Authors:** Le Wei, Matthew N. Ahmadi, Joanna M. Blodgett, Elroy J. Aguiar, Raaj Kishore Biswas, Borja del Bozo Cruz, Emmanuel Stamatakis

## Abstract

**Background:** Research on the health effects of stepping intensity in free-living environments is limited and inconclusive. Inconsistent use of stepping intensity estimation metrics could explain current equivocal results. We aimed to examine and compare a range of different cadence-based metrics in terms of their multivariable-adjusted associations with all-cause (ACM) cardiovascular disease (CVD), cancer and physical-activity (PA)-related cancer mortality.

**Methods:** This prospective cohort study included participants with valid wrist-worn accelerometer data from the UK Biobank. We estimated stepping intensity using ten different cadence-based metrics, including eight peak-cadence metrics (defined as averaged steps / min of the highest but not necessarily consecutive minutes) that most of whom have appeared in prior literature, plus two non-peak-cadence metrics: 1) average daily cadence, defined as steps/accelerometer wearing mins, and 2) average cadence of purposeful steps, defined as averaged steps / min of minutes with ≥40 steps. We rescaled each metric into a standardised cadence scale with mean of 0 and standard deviation (SD) of 1, using (absolute–mean)/SD. We compared the dose-response associations of each stepping intensity estimation metrics with mortality outcomes using previously published modelling involving Cox-restricted-cubic-spline model, presented as overlay plots on standardised and absolute cadence scales.

*Results:* Among 70,336 participants (age [SD], 61.6 [7.8] years; female, 40,933 [58%]) followed up for a median of 8.0 years, all cadence-based metrics, besides the average cadence of purposeful steps, exhibited a comparable beneficial dose-response association with ACM/CVD/cancer mortality, with 95% CI largely overlapped (e.g., at −0.2 standardised steps/min, the hazard ratio (HR) of ACM for peak 1- and peak 30-min cadence were: 0.72, 95%CI [0.65, 0.82] and 0.66 [0.60, 0.73], respectively). The average cadence of purposeful steps only did not show dose-response associations with mortality outcomes (e.g., the HR that corresponds to the standardised median for the average cadence of purposeful steps in ACM was 0.98 [95% CI: 0.86, 1.12].

**Conclusion:** Besides the average cadence of purposeful steps, all stepping intensity estimation metrics demonstrated comparable beneficial dose-response associations with mortality of all-cause, CVD and cancer, suggesting these cadence-based metrics may be used interchangeably for estimating associations of free-living stepping intensity with health outcomes and applied in different research scenarios accordingly.

## Introduction

With the widespread use of wearable devices in recent years, self-monitoring of stepping behaviour and goal setting is easier than ever before^1^. While prior evidence on the potential health benefits associated with step quantity is relatively consistent^2,3^, evidence on intensity, a crucial aspect emphasized in current major physical activity (PA) guidelines^4–6^, remains inconsistent and limited^7–9^. This inconsistency might preclude researchers from making evidence-based conclusions regarding the health benefits of stepping intensity to inform step-based public health guidelines. Current equivocal evidence might be explained by limited and inconsistent estimation metrics of free-living stepping intensity. The stepping activities, as a fundamental component of day-to-day PA^10^ (e.g., walking), should be theoretically associated with intensity-specific health benefits. As such, identifying appropriate stepping intensity estimation metrics through a comprehensive comparison among various metrics is crucial for studies related to free-living steps.

Peak cadence is defined as the average steps/min recorded for the highest but not necessarily consecutive minutes in a day^11,12^, and thus has the advantage of reflecting the health benefits of short bout (e.g., less than 10 mins) PA^13,14^. Peak cadence-based metrics (e.g., peak 30-minute cadence) are shaped by both intensity and consistency of stepping behavior across the measurement period, representing individuals “natural best effort” relative to their capacity in free-living environments^11,15^. Prior evidence on long-term health effects of peak cadence metrics is equivocal. The most commonly used stepping intensity estimation metric, peak 30- and 60-min cadence was inversely associated with lower risk of all-cause mortality (ACM) in a large harmonized meta-analysis, whereas other studies limited to older women^16^ and middle-aged adults^17^ did not exhibit an such association using peak 30-min cadence^14^. Some evidence has also indicated that peak 30-min cadence is associated with lower mortality of CVD and cancer^18^.

The peak 30-minute cadence was initially designed to align with physical activity guidelines recommending 30 minutes of moderate to vigorous physical activity (MVPA) per day for adults. These guidelines are primarily based on self-reported data, which likely results in an overestimation of MVPA minutes^19^. Recent device-based large cohort studies have shown that only 4-5 mins of vigorous-intensity PA (VPA)^20,21^ or roughly 20 mins of MVPA^14^ were associated with 30-40% risk reduction in mortality of all-cause, CVD and cancer^22^, with more MVPA or VPA minutes yielding slightly additional health benefits in diminishing manner. Peak cadence of shorter time interval might be able to capture the health benefits associated with stepping intensity without including excess non-stepping behaviours (e.g., sedentary behaviours) as in that of peak 30- or 60-min cadence, demonstrating potential of being a better estimation metric reflecting the natural best exertion on free-living stepping. Thus. it may be worthwhile to explore the relevance of peak cadence over shorter time intervals (i.e., peak 1, 5-, 10-, 15-, 20-, 25-min cadences) in addition to the commonly used peak 30-min cadence.

Besides peak cadence metrics, one prior study indicates that average cadence, defined as total daily step counts divided by total valid accelerometer wear minutes, was inversely associated with ACM risk in older adults^23^, showing its potential as a cadence-based metric estimating stepping intensity. Meanwhile, minutes spent at a cadence ≥40 steps/min (i.e., purposeful steps^24,25^) has also exhibited a beneficial association with ACM^16^. However, there has been no study investigating the potential of the average cadence of purposeful steps.

Accordingly, we explored and compared the dose-response associations of ten different stepping-intensity estimation metrics, including eight peak-cadence metrics and two other cadence-based metrics, with mortality of all-cause, CVD, cancer and PA-related cancer, in a large prospective cohort of UK adults.

## Methods

### Participants

The UK Biobank is a large prospective cohort with 502,616 UK adults aged 40-69 years recruited between 2006 and 2010^26^. Participants completed baseline measurements and provided written consent to use their data. The ethical approval was provided by the UK National Health Service, National Research Ethics Service (Ref 11/NW/0382).

From 2013 to 2015, 103,684 UK biobank participants were mailed and wore the Axivity AX3 (Axivity Ltd, Newcastle Upon Tyne, United Kingdom) wrist-worn triaxial accelerometer on their dominant wrist for 7 days continuously. The AX3 was initialised to capture triaxial acceleration data at a sampling frequency of 100 Hz and a dynamic range of ±8g. A monitoring day was considered valid if the wear time exceeded 16 hours^25^. Participants returned the devices by mail and the data were calibrated and non-wear periods were identified^27,28^. Participants needed at least three valid monitoring days, including a minimum of one weekend day to be included^18,29^. We excluded those with insufficient valid wear time, missing covariate data, prevalent CVD or cancer history (ascertained through hospital admission records), poor self-reported health, and death within one year after PA assessment^16,18^. We calculated steps during periods of ambulation using a tuned signal peak detection method^30^ with step detection accuracy of 89%, total steps mean absolute percent error of 10%^31^, and a mean bias of 9%^32^

### Definition of stepping intensity cadence-based estimation metrics

#### Peak-cadence metrics: peak 1-, 5-, 10-, 15-, 20-, 25-, 30-, 60-min cadence

Minute-by-minute step data were rank-ordered from the highest to lowest^11^ for each valid accelerometer wear day. Then, we selected the highest steps/min for 1, 5, 10, 15, 20, 25, 30, and 60 minutes (not necessarily consecutive minutes), and calculated the average steps/min over the corresponding time interval for each valid wear day. Finally, we averaged them over the total number of valid wear days.

#### Other cadence-based metrics: average cadence, average cadence of purposeful steps

To calculate average cadence, we divided the total step counts by the total valid accelerometer wear minutes to calculate the steps per minute for each valid wear day. Then we calculated the mean over the total valid wear days^23^. To calculate the average cadence of purposeful steps, we first identified minutes where participants took at least 40 consecutive steps. For each valid wear day, we summed the total step count during these minutes and divided it by the total minutes of the steps. The average cadence of purposeful steps was then calculated by averaging these daily values across all valid wear days.

### Outcome ascertainment

Participants were followed up to 30 November 2022, with deaths obtained through linkage with the National Health Service (NHS) Digital of England and Wales or the National Records of Scotland. Based on ICD-10 codes from both primary and contributory death cause, we defined CVD mortality as death from diseases of the circulatory system (ICD-10 codes: I0, I11, I13, I20–I51, I60–I69), excluding hypertension and diseases of arteries and lymph^33^. We defined cancer mortality as death attributed to any cancer excluded in situ, benign, uncertain, non-melanoma skin cancers, or non-well-defined cancers (ICD-10 codes: C0-C6, C70-C75, C7A, C8, C9)^34^. PA-related cancer mortality was defined as death from 13 site-specific cancers associated with low PA: bladder, breast, colon, endometrial, oesophageal, adenocarcinoma, gastric cardia, head and neck, kidney, liver, lung, myeloid leukemia, myeloma, and rectal (ICD-10 codes: C0-C3, C5-C9, C40-C42, C45-C49)^18^.

### Covariates

Based on similar peer-reviewed literature examining the association between stepping intensity and mortality^16–18^, all analyses were adjusted for age, sex, ethnicity, valid accelerometer wear days, smoking status, alcohol consumption, sleep duration, sedentary time, fruit and vegetable consumption, education level, economic status, family history of CVD and cancer, and medication use of cholesterol, blood pressure and diabetes, and daily step counts using the residual method^2,35^.

### Statistical analysis

We excluded data below 1st and above 99th percentile of the distribution across cadence-based metrics to minimise the influence of sparse data^18^. We standardised the cadence values, calculated as (absolute – mean)/standard deviation^36,37^, across cadence-based metrics to rescale them for cross comparison. Each metric was centered around a mean of 0 with a standard deviation (SD) of 1. We assessed time-to-event dose-response associations of standardised stepping intensity with ACM using cox restricted cubic spline model. We placed knots at 6th, 34th, and 67th percentile of the exposures’ distributions as they right skewed. We set the reference level at the 5^th^ percentile^34^. For cause-specific outcomes, we used the Fine and Grey model to account for competing risks^18,38^. We assessed proportional hazard assumptions through Schoenfeld residuals and observed no violation. We presented overlay dose-response plots on a scale of standardised cadence to visually compare the dose-response associations of similar proportion of stepping intensity across metrics were compared in a same scale^37^, better informing which metric demonstrated a more pronounced association with mortality. For those comparisons, we assessed differences in effect size at the standardised cadence of −0.2, −0.5 for lower scale, and 3.0 and 3.3 for upper scale. To examine the differences in doses and corresponding health effects of important indices between metrics, we assessed the minimum effect dose corresponds to the least discernible useful effect^39^ (i.e., 50% of the optimal risk reduction in PA related studies)^20^ and the maximum useful dose that corresponds to the optimal risk reduction (nadir of the curve)^20^. Meanwhile, we assessed the median and the corresponding health effect across stepping intensity estimation metrics^40^. We presented the distribution of each metric to examine the appropriateness of using standarisation scaling. We repeated above analysis using absolute cadence.

We assessed the robustness of our findings with four sensitivity analysis: First, we presented overlay dose-response plots on the scale using normalised cadence, an alternative to the standardized cadence calculated as (cadence - minimum cadence) / (maximum cadence - minimum cadence) resulting in the same normalised cadence range across all metrics from 0 to 1^36^; Second, to further reduce the influence of outliers on the dose-response curve, we provided the overlay dose-response plots excluding data below 2.5^th^ and above 97.5^th^ percentile of the exposure distribution; Finally, we examined the dose-response associations with knots placing at 10^th^, 50^th^ and 90^th^ percentile of standardised cadence distribution^20^. We performed all analyses using *R* statistical software (version 4.2.2).

## Results

### Description of the study sample

Our sample included 70,336 participants (mean [SD] age, 61.6 [7.8] years; female, 40,933 [58%]) followed up for a median of 8.0 years (SD: 0.9) with 2,037 mortalities (CVD: 553; cancer: 1209; PA-cancer: 409) (Supplementary Table 1 and flowchart in Supplementary Fig 1). The distributions of standardised stepping intensity estimation metrics were comparable (Supplementary Fig.2b and 2b).

### Association of stepping intensity with all-cause mortality

We observed a steeper gradient of the dose-response associations between the peak cadence of longer intervals and ACM compared to that of shorter intervals at the lower scale of standardised cadence, with 95% CI largely overlapping (Fig. 1). For example, at −0.2 standardised steps/min, the HR corresponds to peak 30- and 1-min cadence was: 0.66 (95% CI: 0.60, 0.73) and 0.72 (0.65, 0.82), respectively (Table 1). However, less steep dose-response associations were shown for the peak cadence of longer intervals at the upper end of standardized cadence scale (Fig. 1), e.g., HR for peak 60- and 5-min at 3.3 standardised cadence was: 0.75 (95% CI: 0.58, 0.96) and 0.67 (0.52, 0.85), respectively (Table 1). The standardised median, minimum dose and the corresponding effect size for ACM were comparable across all peak cadence metrics, e.g., the minimum dose for peak 30- and 15-min cadence metrics were –0.97 and −0.95, respectively, with corresponding HR of 0.83 (95% CI: 0.79, 0.88) and 0.83 (0.78, 0.88), respectively. The dose-response association of the average cadence with ACM was similar to that of peak 30-min cadence showing a steep gradient, whereas the average cadence of purposeful steps exhibited no dose-response relation (Fig.2). For example, standardised median for the average cadence and the average cadence of purposeful steps were: 0.63, and 0.59, respectively, with corresponding HR of 0.69 (95% CI: 0.60, 0.78) and 0.98 (0.86, 1.12), respectively.

**Fig. 1.**
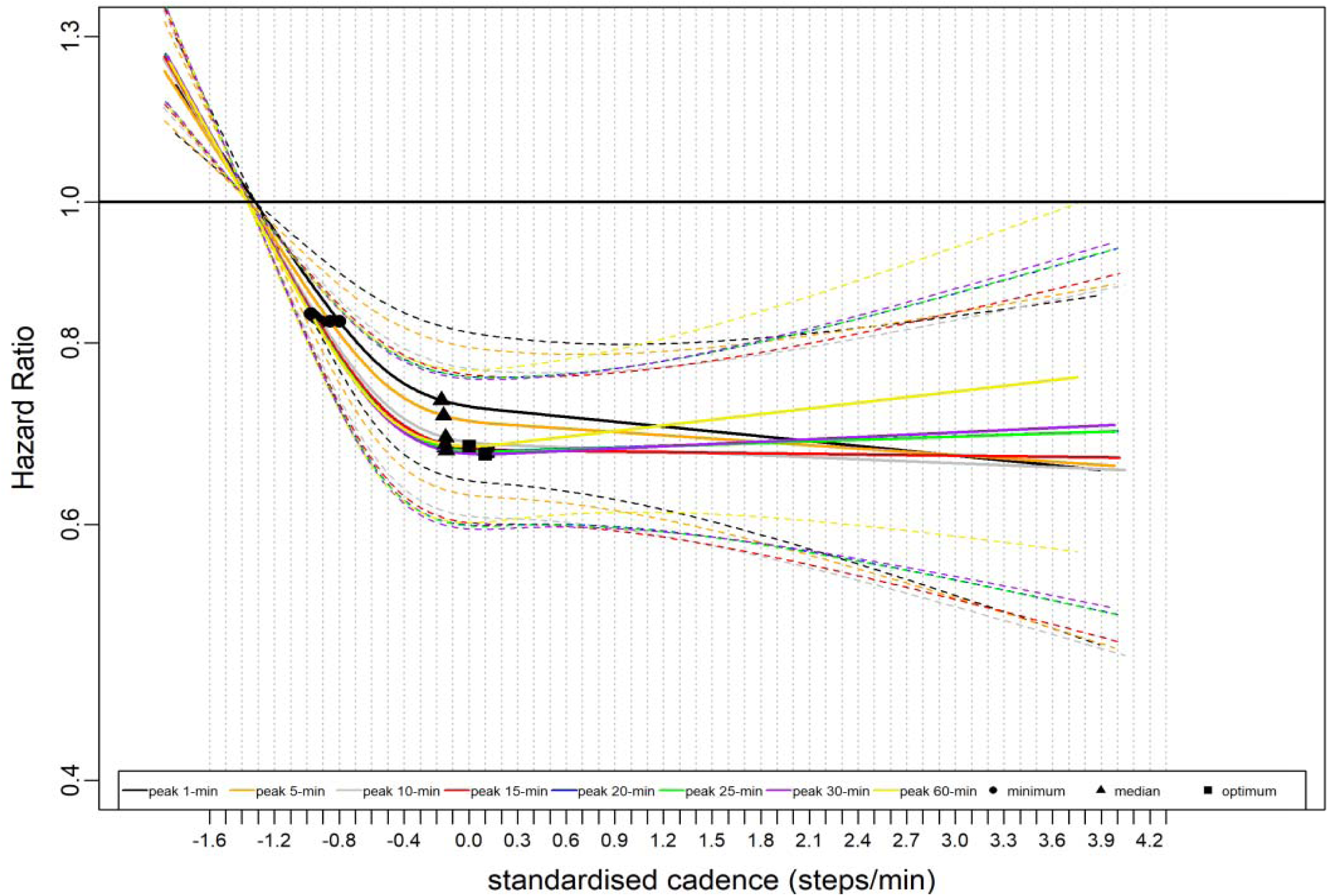
Dose-Response Association of Standardised Stepping Intensity Estimated across Peak Cadence Metrics with All-Cause Mortality Total sample size is 70,174. The events for peak 1-min cadence are 2,029; peak 5-min cadence, 2,023; peak 10-min cadence, 2,019; peak 15-min cadence, 2,021; peak 20-min cadence, 2,022; peak 25-min cadence, 2,020; peak 30-min cadence, 2,021; peak 60-min cadence, 2,029. The standardised cadence was calculated as ([peak cadence - mean] / standard deviation). The circle indicates the ED50 value i.e., minimum, the standardised cadence that associated with 50% of the optimal risk reduction; The triangle indicates the median standardised cadence. The square indicates the standardised cadence that associated with optimal mortality risk reduction (Note: the square was not annotated if there was no nadir point). We analysed the dose-response associations using cox-regression model and adjusted for age, sex, accelerometer wearing duration, average daily steps, smoking status, alcohol consumption, sleep duration, Townsend deprivation score, sedentary time, education levels, self-reported parental history of CVD and cancer, and self-reported medication use (cholesterol, blood pressure, and diabetes). The reference level was set as the 5^th^ percentile of each peak cadence metric.

**Fig. 2.**
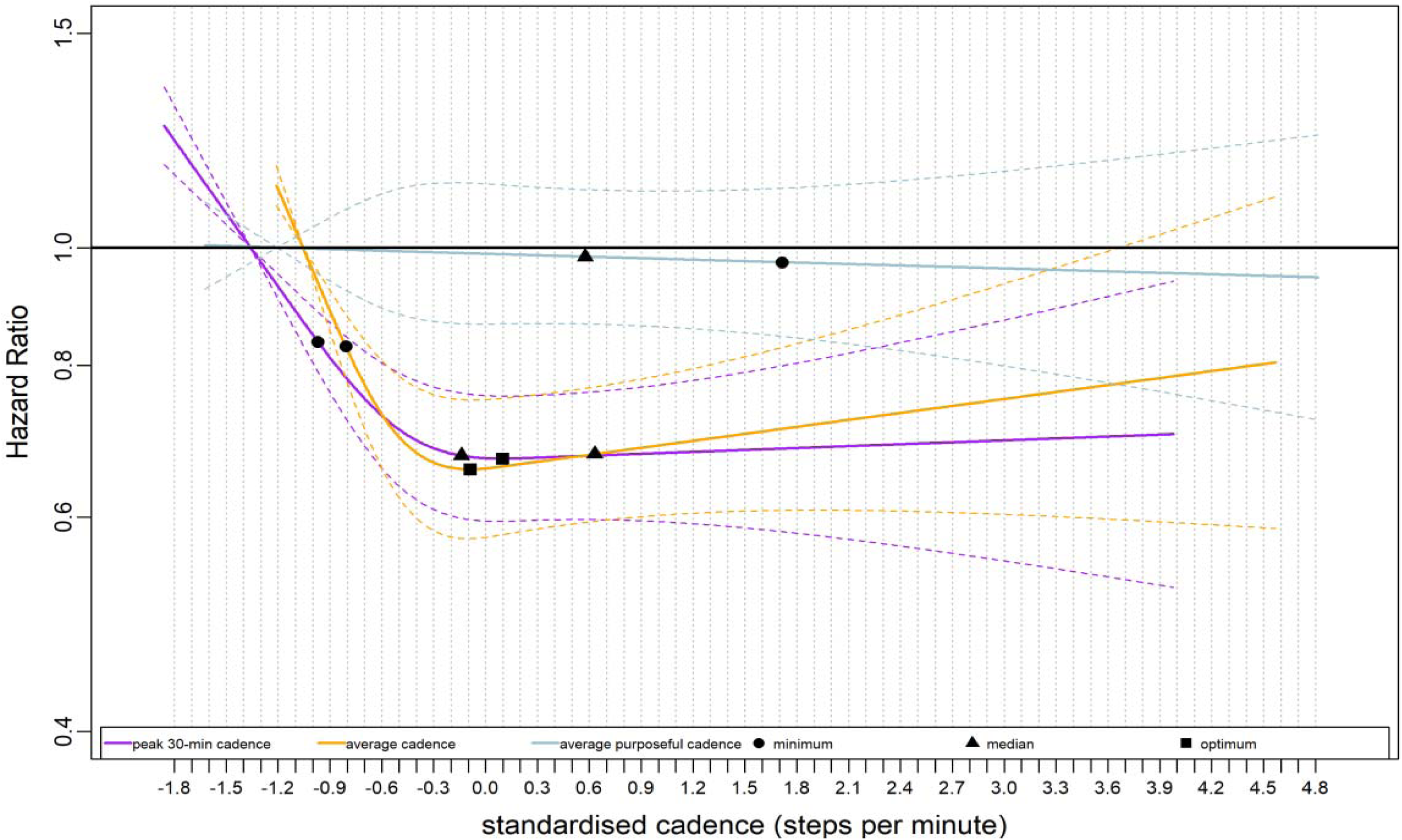
Dose-Response Association of Standardised Stepping Intensity Estimated across Non-peak cadence metrics (Average Cadence, Average Cadence of Purposeful Steps) and Peak 30-min Cadence with All-Cause Mortality We compared two non-peak cadence metrics and a representative peak cadence metric (peak 30-min cadence) in this figure. Total sample size is 70,174. Events for the average cadence per day is 2,022; average cadence of purposeful steps per day, 2,064; peak 30-min cadence, 2,021; The standardised cadence was calculated as ([exposure - mean] / standard deviation). The circle indicates the ED50 value i.e., minimum, the standardised cadence that associated with 50% of the optimal risk reduction; The triangle indicates the median standardised cadence. The square indicates the standardised cadence that associated with optimal mortality risk reduction (Note: the square was not annotated if there was no nadir point). We analysed the dose-response associations using cox-regression model and adjusted for age, sex, accelerometer wearing duration, average daily steps, smoking status, alcohol consumption, sleep duration, Townsend deprivation score, sedentary time, education levels, self-reported parental history of CVD and cancer, and self-reported medication use (cholesterol, blood pressure, and diabetes). The reference level was set as the 5^th^ percentile of each stepping intensity metric.

### Association of stepping intensity with CVD mortality

The gradient pattern of the dose-response associations for CVD mortality was similar to those observed in ACM (Fig 3), with peak cadence of longer intervals showing a steeper gradient at the lower scale of the standardised cadence and a less steep gradient at the upper scale compared to those of short intervals. For example, the HR for CVD mortality for peak 5- and 30-min cadence at −0.2 standardised cadence was: 0.61 (95% CI: 0.49, 0.75) and 0.57 (0.46, 0.71), respectively; and at 3.3 standardised cadence: 0.61 (0.38, 0.98) and 0.65 (0.40, 1.04), respectively (Table 2). The standardised median, minimum and maximum dose and the corresponding effect size were comparable across peak cadence metrics, e.g., the standardised minimum doses for peak 5- and peak 30-min cadence were −0.97, and −1.00, respectively, with corresponding HR of 0.80 (95% CI: 0.72, 0.89) and 0.79 (0.71, 0.87), respectively. The dose-response association of the standardised average cadence with CVD mortality was similar to that of peak 30-min cadence with a steep gradient, whereas the average cadence of purposeful steps showed no dose-response association (Fig. 4). For example, the standardised median for the average cadence and the average cadence of purposeful steps were: 0.69 and 0.65 respectively, but with different corresponding HR of 0.60 (95% CI: 0.47, 0.77) and 0.99 (0.77, 1.26), respectively (Table 2).

**Fig. 3.**
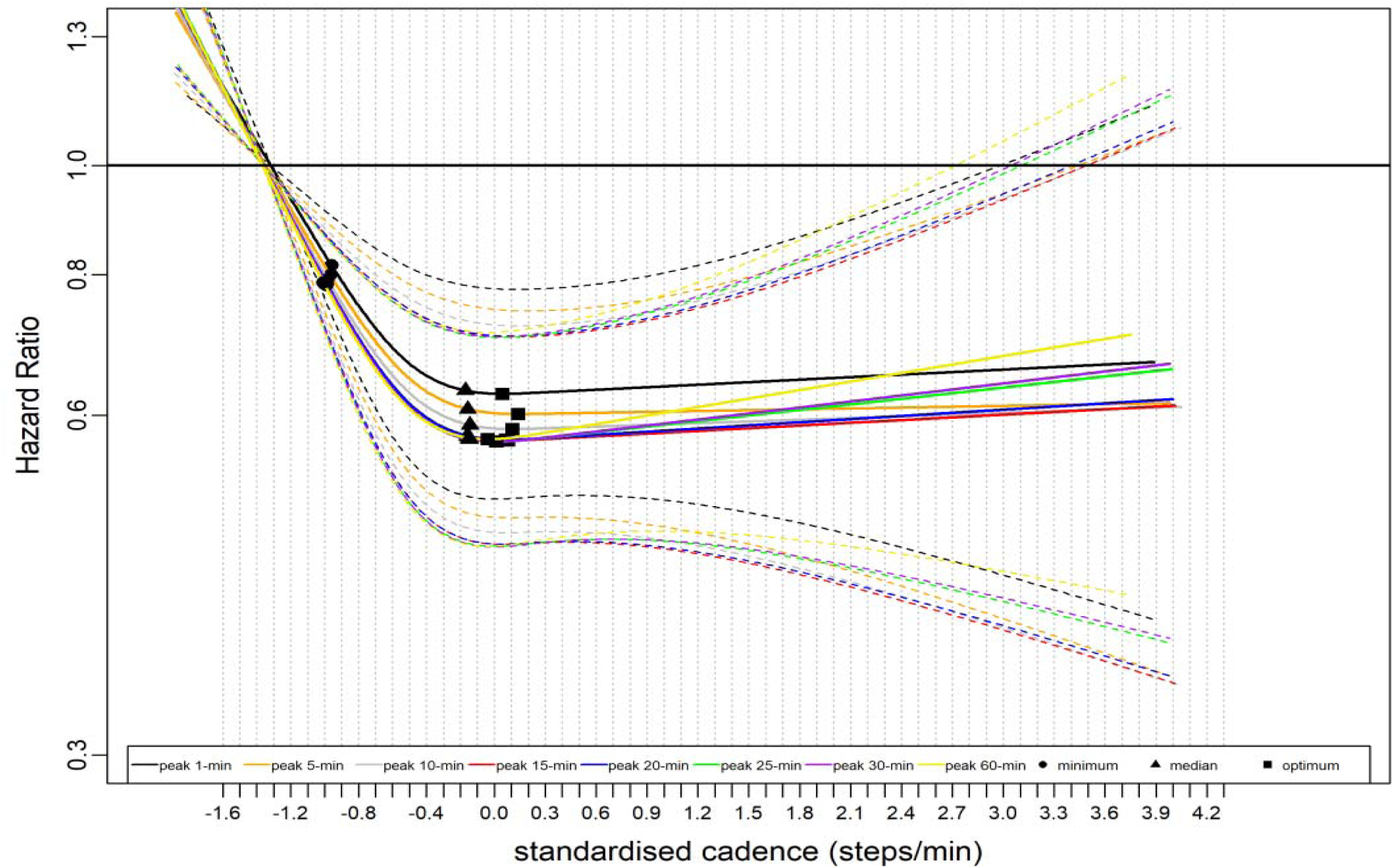
Dose-Response Association of Standardised Stepping Intensity Estimated across Peak Cadence metrics with CVD Mortality Total sample size is 70,174. The events for peak 1-min cadence are 549; peak 5-min cadence, 547; peak 10-min cadence, 546; peak 15-min cadence, 546; peak 20-min cadence, 546; peak 25-min cadence, 546; peak 30-min cadence, 548; peak 60-min cadence, 553; The standardised cadence was calculated as ([exposure - mean] / standard deviation). The circle indicates the ED50 value i.e., minimum, the minimal cadence associated with 50% of the optimal risk reduction; The triangle indicates the standardised median cadence. The square indicates the standardised cadence that associated with optimal mortality risk reduction (Note: the square was not annotated if there was no nadir point). We used Fine and Grey model to analyse the dose-response association and adjusted for age, sex, accelerometer wearing duration, average daily steps, smoking status, alcohol consumption, sleep duration, Townsend deprivation score, sedentary time, education levels, self-reported parental history of CVD and cancer, and self-reported medication use (cholesterol, blood pressure, and diabetes). The reference level is 5^th^ percentile of each standardised peak cadence metric.

**Fig. 4.**
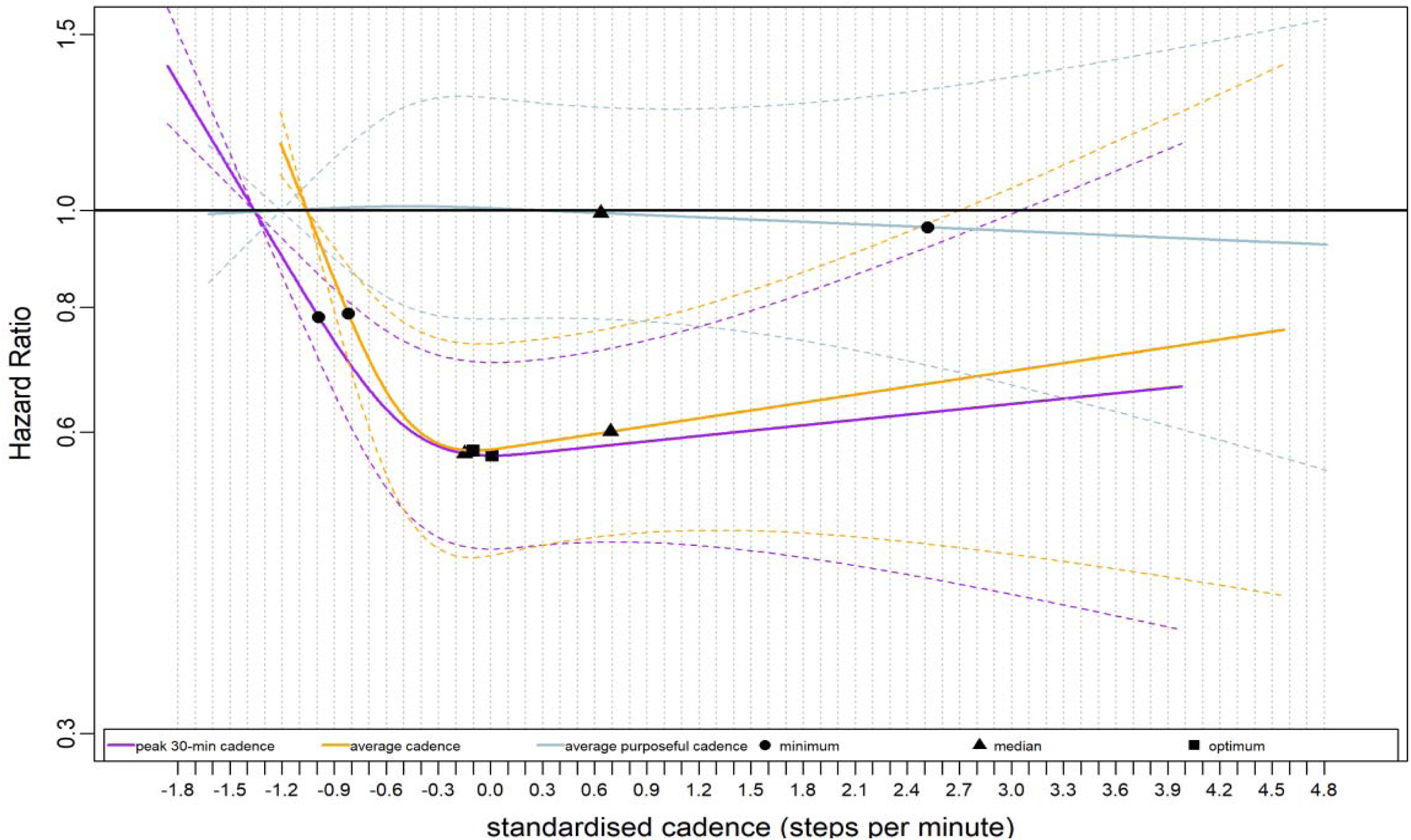
Dose-Response Association of Standardised Stepping Intensity Estimated by the Average Cadence, Average Cadence of Purposeful Steps, and Peak 30-min Cadence with CVD Mortality We compared two non-peak cadence metrics and a representative peak cadence metric (peak 30-min cadence) in this figure. Total sample size is 70,174. The events for the average cadence is 545; average cadence of purposeful steps, 569; peak 30-min cadence, 548; The standardised cadence was calculated as ([exposure - mean] / standard deviation). The circle indicates the ED50 value i.e., minimum, the standardised cadence that associated with 50% of the optimal risk reduction; The triangle indicates the median standardised cadence. The square indicates the standardised cadence that associated with optimal mortality risk reduction (Note: the square was not annotated if there was no nadir point). We used Fine and Grey model to analyse the dose-response association and adjusted for age, sex, accelerometer wearing duration, average daily steps, smoking status, alcohol consumption, sleep duration, Townsend deprivation score, sedentary time, education levels, self-reported parental history of CVD and cancer, and self-reported medication use (cholesterol, blood pressure, and diabetes). The reference level is 5^th^ percentile of each standardised peak cadence metric.

### Association of stepping intensity with cancer mortality

A modest difference was observed for the dose-response associations with cancer mortality across peak cadence metrics (Fig 5). E.g., at 3.0 standardised steps/min, the corresponding HR for peak 1- and peak 60-min cadence were 0.76 (95% CI: 0.57, 1.01) and 0.82 (0.61, 1.12), respectively (Table 3). The standardised median, minimum and maximum dose and the corresponding effect size for cancer mortality were comparable across peak cadence metrics, e.g., the standardised minimum dose for peak 5- and peak 30-min cadence were 0.73 and 0.48, respectively, with comparable corresponding HR of 0.86 (95% CI: 0.72, 1.00) and 0.86 (0.72, 1.01), respectively. The dose-response association of the standardised average cadence was steeper than that of the average cadence of purposeful steps (Fig 6).

**Fig. 5.**
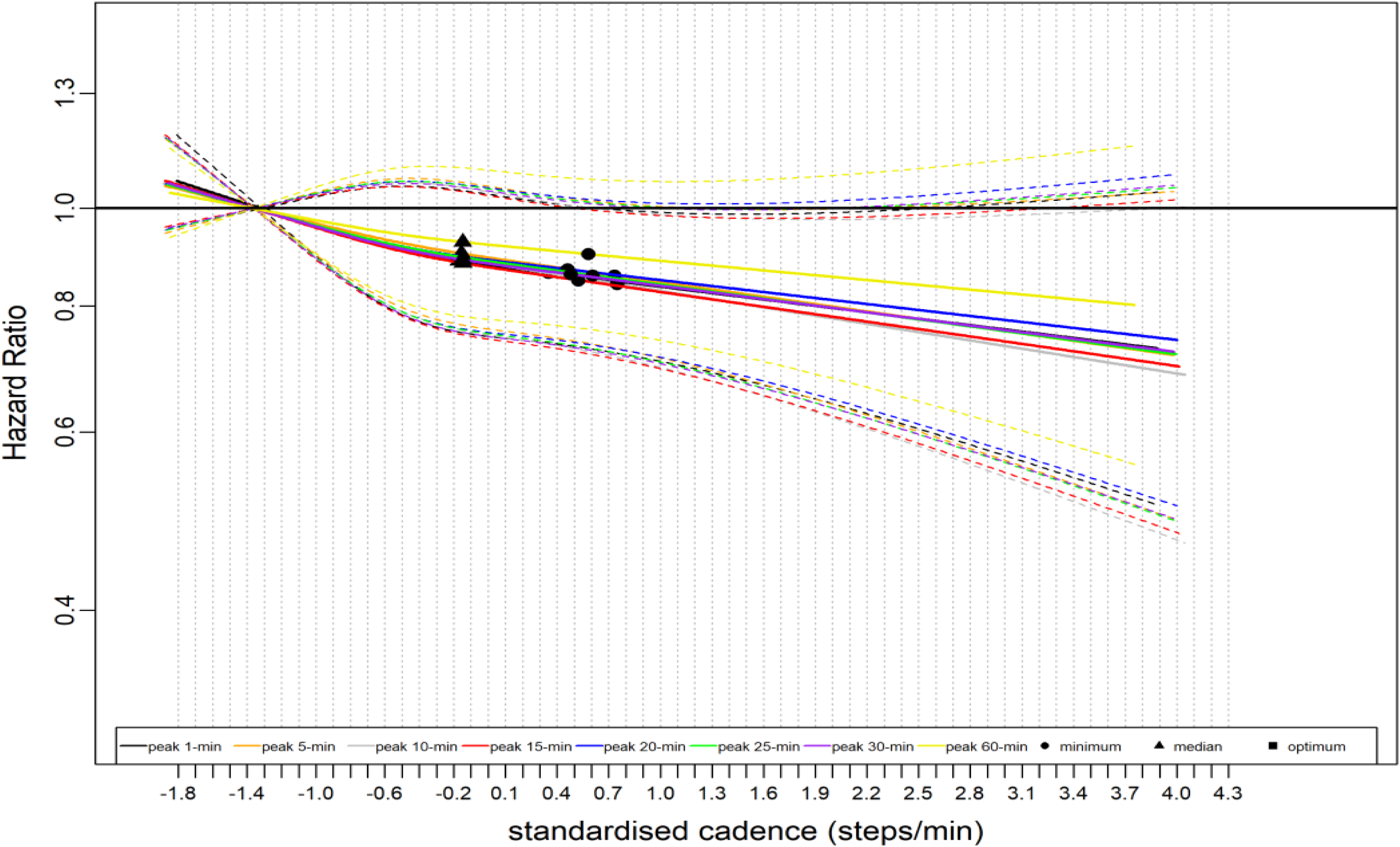
Dose-Response Association of Standardised Stepping Intensity Estimated across Peak Cadence Metrics with Cancer Mortality Total sample size is 70,174. The events for peak 1-min cadence is 1,205; peak 5-min cadence, 1,204; peak 10-min cadence, 1,201; peak 15-min cadence,1,201; peak 20-min cadence, 1,203; peak 25-min cadence, 1,201; peak 30-min cadence, 1,201; peak 60-min cadence, 1,201. The standardised cadence was calculated as ([exposure - mean] / standard deviation). The circle indicates the ED50 value i.e., minimum, the minimal steps per min associated with 50% of the optimal risk reduction; The triangle indicates the median steps per min. The square indicates the standardised cadence that associated with optimal mortality risk reduction (Note: the square was not annotated if there was no nadir point). We used Fine and Grey model to analyse the dose-response association and adjusted for age, sex, accelerometer wearing duration, average daily steps, smoking status, alcohol consumption, sleep duration, Townsend deprivation score, sedentary time, education levels, self-reported parental history of CVD and cancer, and self-reported medication use (cholesterol, blood pressure, and diabetes). The reference level is 5^th^ percentile of each standardised peak cadence metric.

**Fig. 6.**
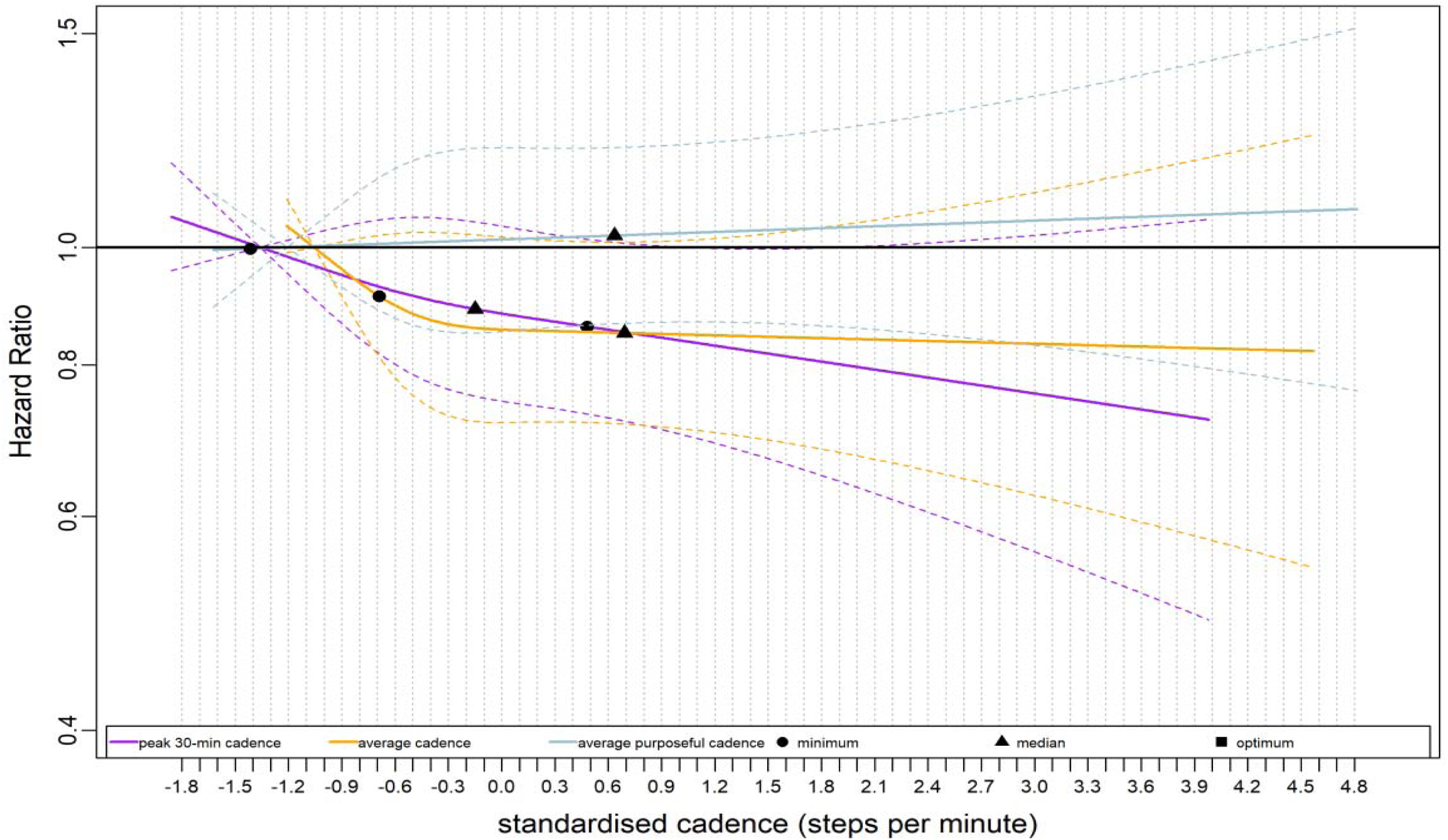
Dose-Response Association of Standardised Stepping Intensity Estimated by the Average Cadence, Average Cadence of Purposeful Steps, and Peak 30-min Cadence with Cancer Mortality We compared two non-peak cadence metrics and a representative peak cadence metric (peak 30-min cadence) in this figure. Total sample size is 70,174. The events for peak 30-min cadence are 1201; average cadence per day, 1205; average cadence of purposeful steps per day, 1214. The standardised cadence was calculated as ([exposure - mean] / standard deviation). The circle indicates the ED50 value i.e., minimum, the standardised cadence that associated with 50% of the optimal risk reduction; The triangle indicates the median standardised cadence. The square indicates the standardised cadence that associated with optimal mortality risk reduction (Note: the square was not annotated if there was no nadir point). We used Fine and Grey model to analyse the dose-response association and adjusted for age, sex, accelerometer wearing duration, average daily steps, smoking status, alcohol consumption, sleep duration, Townsend deprivation score, sedentary time, education levels, self-reported parental history of CVD and cancer, and self-reported medication use (cholesterol, blood pressure, and diabetes). The reference level is 5^th^ percentile of each standardised peak cadence metric.

### Association of stepping intensity with physical activity related cancer mortality

Peak 60-min cadence metric showed less pronounced dose-response association comparing to those of other peak cadence metrics that exhibited similar associations (Supplementary Fig. 4). The standardised average cadence and the average cadence of purposeful steps showed comparable dose-response association with PA-related cancer mortality (Supplementary Fig. 5).

### Association of absolute stepping intensity with outcomes

Supplementary Fig. 6-13 demonstrated the dose-response associations of stepping intensity, estimated using the absolute cadence, with mortality outcomes. All dose-response associations were comparable to those using standardized cadence in terms of effect size, with sizeable differences in absolute cadence showing that the longer the peak cadence interval, the more the dose-response curve shifted towards the lower end of the absolute cadence scale. For example, the median and HR for peak 60- and peak 1-min cadence were 54.9 steps/min, 0.68 (95% CI: 0.60, 0.77) and 147.9 steps/min, 0.75 (0.67, 0.84) for ACM.

### Sensitivity analysis

Placing knots at the 10^th^, 50^th^, and 90^th^ percentile of the exposure distribution (Supplementary Fig.14) or using the normalised cadence scale (Supplementary Fig. 15) or excluding outliers below 2.5^th^ and above 97.5^th^ percentile of the exposure distribution (Supplementary Fig.16) did not materially change the results observed in the main analysis.

## Discussion

Despite the importance of PA intensity to public health, to date, the evidence on the health effects of stepping intensity in free-living environments is not established. One possible explanation is the inconsistent use of different stepping intensity estimation metrics. To our knowledge, this study is the first to compare the dose-response associations of various stepping intensity estimation metrics with long-term health outcomes. The peak cadence and the average cadence metrics revealed comparable beneficial dose-response associations with mortality of all-cause, CVD and cancer across the spectrum of the standardised cadence scale, suggesting that these stepping intensity estimation metrics may be used interchangeably to estimate long-term health benefits of different stepping intensity levels in free-living environments. However, the average cadence of purposeful steps did not demonstrate beneficial associations with mortality of all-cause, CVD, cancer and PA-related cancer.

The dose-response curves and corresponding effect size across all peak cadence and average cadence metrics showed substantial overlap. Crucial point estimates, such as the least discernable useful effect, the maximum mortality risk reduction, and their corresponding standardised stepping intensity were similar^26^.

Prior evidence has consistently shown the substantial health benefits of vigorous-intensity PA (VPA)^41,42^. As few as roughly 5 VPA minutes per day, primarily consisting of stepping activities, were associated with a 30-40% lower HR accounting for the majority of the risk reduction in all-cause and CVD mortality compared to those who rarely engage in VPA^20,21^. For physically active individuals who might accumulate more mins of higher intensity stepping activities (e.g., jogging), peak cadence of time interval shorter than 30 mins might reflect a similar mortality risk reduction to those of longer interval, resulting in comparable dose-response associations across peak cadence metrics. For physically less active adults with a limited amount of higher intensity stepping mins, peak cadence of shorter time intervals might be sufficient to reflect the mortality risk reduction as those of longer intervals. Notably, the peak 1-min cadence also demonstrated a standardised dose-response association comparable to the other peak cadence metrics, suggesting that even the fastest walking minute one each day conveys useful information. In summary, beneficial and comparable dose-response associations with mortality risk were revealed for middle-aged and older adults, regardless of the length of time interval in peak cadence metrics.

Albeit higher amount of daily purposeful steps was associated with potential health benefits for middle-aged and older adults^18,43^, we did not observe a dose-response association for the average cadence of purposeful steps with mortality. This could be partly explained by the variation in values of denominators (i.e., the number of minutes above 40 steps/min) used in calculation for different participants. For those adults with higher amount of daily purposeful steps, a corresponding higher minute spent on purposeful steps might result in similar averaged cadence of purposeful steps to others with lower purposeful steps and mins. As such, the average cadence of purposeful steps was unable to reflect the mortality risk reduction demonstrated in higher daily purposeful step counts, resulting in different dose-response associations with other cadence-based metrics using fixed denominators (e.g. 30 minutes for all participants when calculating peak 30-min cadence metric).

The peak cadence of longer intervals and the average cadence might be more likely to involve mins with zero or near-zero steps in their calculations than those of shorter ones, resulting in less representative of “stepping minutes”. For instance, many mins of near-zero steps were included in the calculation of peak 30- and 60-min cadence as the average daily stepping time in our sample were roughly 20 mins. From the perspective of representing the intensity of free-living stepping, peak cadence metrics using shorter time-interval might be more appropriate than those of longer ones. Shorter interval peak cadence metrics, such as the peak 1- and 5-min cadence, showed the advantage of having a lower correlation with daily step counts. Since these metrics yield similar mortality risk reduction to those from longer intervals, applying peak cadence of shorter intervals might attenuate the confounding effect of daily step counts in research scenarios examining independent health benefit of free-living stepping intensity.

Our study has several strengths. We examined a comprehensive set of stepping intensity estimation metrics in our studies. Peak cadence and average cadence-based metrics have the advantage of reflecting health benefits of free-living stepping intensity across diverse groups. We conducted four different sensitivity analyses including one using an alternative scaling method to assess robustness of our conclusions. Our study used large prospective cohort with average of almost 8 years of follow-up. In addition, stepping intensity was estimated based on accelerometer data collected during a week, and may not reflect the change of stepping intensity pattern over time. Accelerometer-measured steps may include “error steps” owing to random wrist movement^44^, but equally may not detect some ambulatory behaviour when wrist movement is not pronounced^45^. Applying minute-level cadence-based metrics might downgrade the high intensity stepping bursts to lower intensity levels, particularly considering that the brief and sporadic stepping bouts are typical in adults’ daily life^46^.

In conclusion, the average cadence of purposeful steps might be inappropriate to estimate free-living stepping intensity. Peak cadence and the average cadence metrics revealed comparable beneficial dose-response associations with mortality of all-cause, CVD and cancer across the spectrum of the standardised cadence scale, suggesting those stepping intensity estimation metrics may be used interchangeably for estimating long-term health effects of stepping intensity in free-living environments, and these metrics may be applied accordingly based on different research scenarios.

## Data Availability

Bona fide researchers can register and apply to use the UK Biobank dataset at http://ukbiobank.ac.uk/register-apply/

