## supplementary files for "Does the choice of stepping intensity metric influence dose-response associations with mortality? A UK population cohort study of 70,174 adults"

Participants who wore accelerometer

(**n=103,684**)

Participants without valid wear time (<2 weekdays and <1 weekend day)

(**n=12,846**)

1. Participants with prevalent CVD (**n=7,605**)
2. Participants with prevalent cancer (**n=7,243**)
3. Death within the 1st year after follow up (**n=120)**

PA-cancer mortality

Removing sparse data (1st and 99th percentile): **(n=70,336)**

Peak cadence metrics:

Peak 1-min: 406

Peak 5-min: 406

Peak 10-min: 407

Peak 15-min: 407

Peak 20-min: 408

Peak 25-min: 408

Peak 30-min: 409

Peak 60-min: 410

Non-peak cadence metrics:

Average cadence: 410

Purposeful cadence: 416

Cancer mortality

Removing sparse data (1st and 99th percentile): **(n= 70,336)**

Peak cadence metrics:

Peak 1-min: 1,210

Peak 5-min: 1,206

Peak 10-min: 1,206

Peak 15-min: 1,208

Peak 20-min: 1,208

Peak 25-min: 1,208

Peak 30-min: 1,209

Peak 60-min: 1,210

Non-peak cadence metrics:

Average cadence:1,209

Purposeful cadence:1,228

Participants with missing covariate data

ethnicity = 298; Townsend deprivation score = 309; smoking=175; alcohol=218; qualification=854; sleep=16; meds = 328; fruit and vegetables = 1,149; poor self-rated health =112;

CVD mortality

Removing sparse data (1st and 99th percentile): **(n= 70,336)**

Peak cadence metrics:

Peak 1-min: 550

Peak 5-min: 550

Peak 10-min: 551

Peak 15-min: 552

Peak 20-min: 552

Peak 25-min: 553

Peak 30-min: 553

Peak 60-min: 557

Non-peak cadence metrics:

Average cadence: 549

Purposeful cadence:571

All-cause mortality

Removing sparse data (1st and 99th percentile): **(n=70,336)**

Peak cadence metrics:

Peak 1-min: 2,037

Peak 5-min: 2,031

Peak 10-min: 2,033

Peak 15-min: 2,034

Peak 20-min: 2,036

Peak 25-min: 2,036

Peak 30-min: 2,037

Peak 60-min: 2,044

Non-peak cadence metrics:

Average cadence: 2,034

Purposeful cadence: 2,082

Sample for all-cause, CVD, cancer and PA-related cancer mortality **(n=71,770)**

(**n=72,170)**

**Supplemental fig 1** Flow diagram of participants

Participants with ≥ 3 days of valid wear time

(**n=94,293)**

Participants with complete covariate data

(**n=86,748)**

**Supplementary Fig. 2 The Distribution of the Absolute and Standardised Peak Cadence Metrics**

**a)**
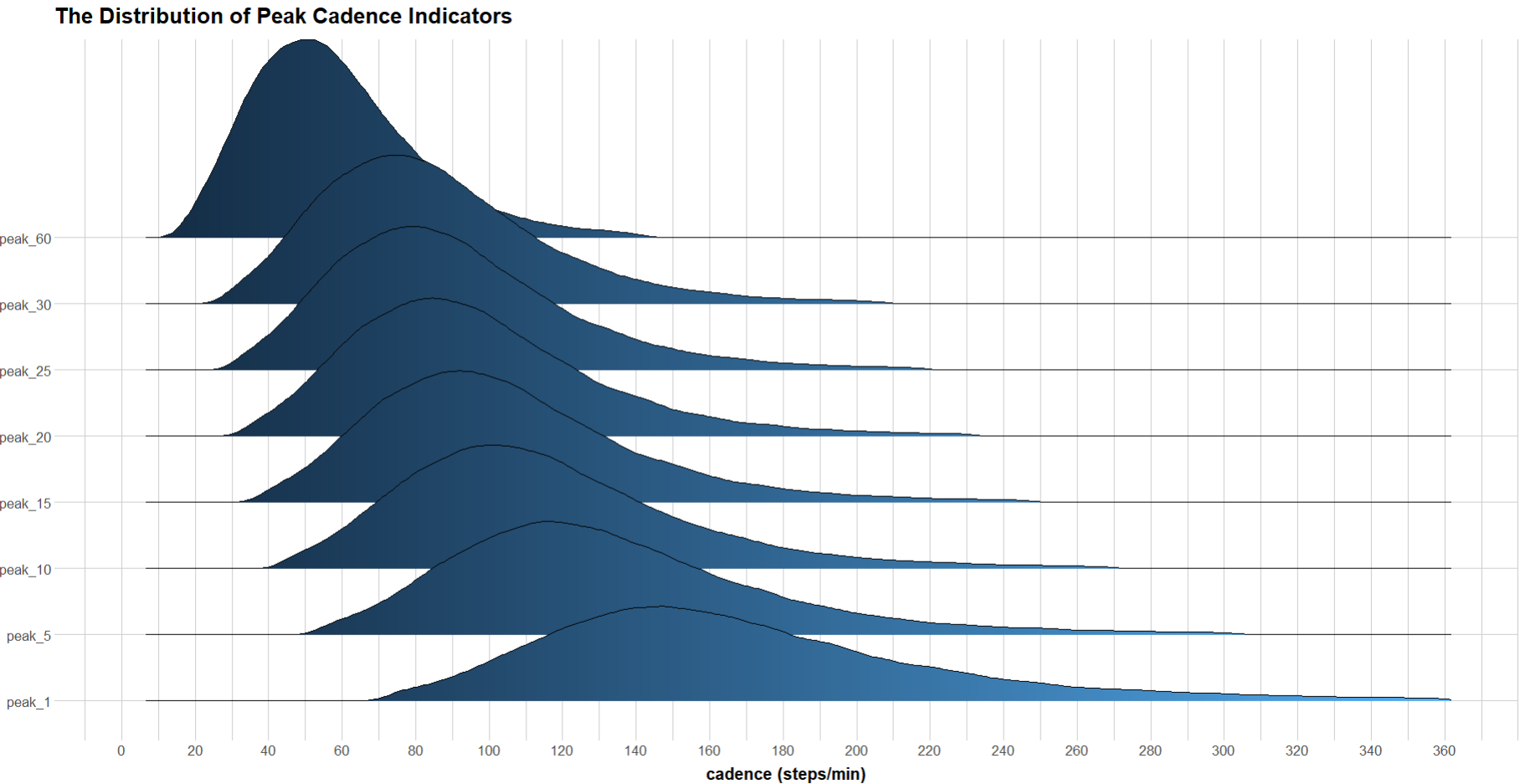

**b)**

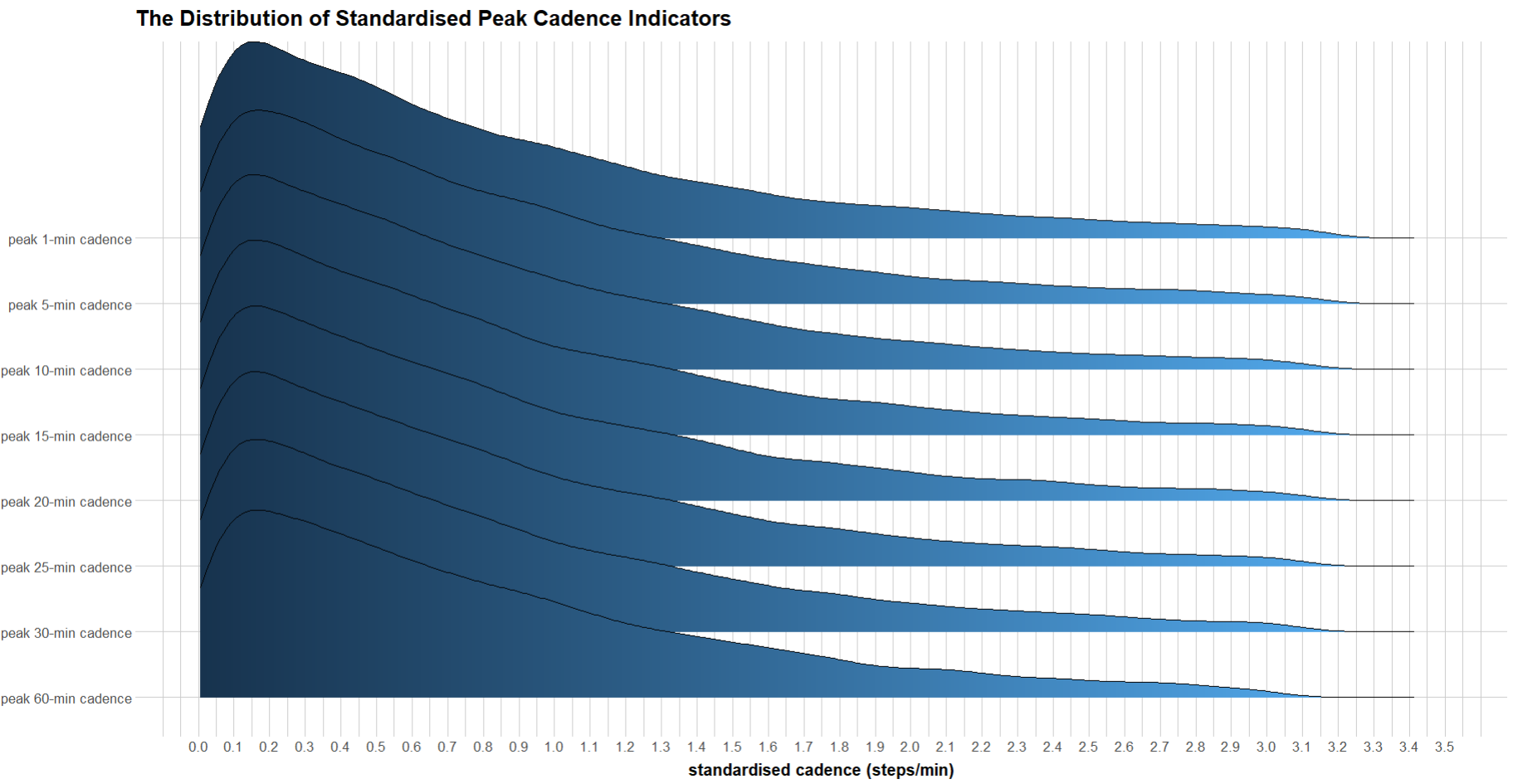

**Supplementary Fig. 3 The Distribution of Absolute and Standardised Non-peak Cadence Metrics**

**a)**

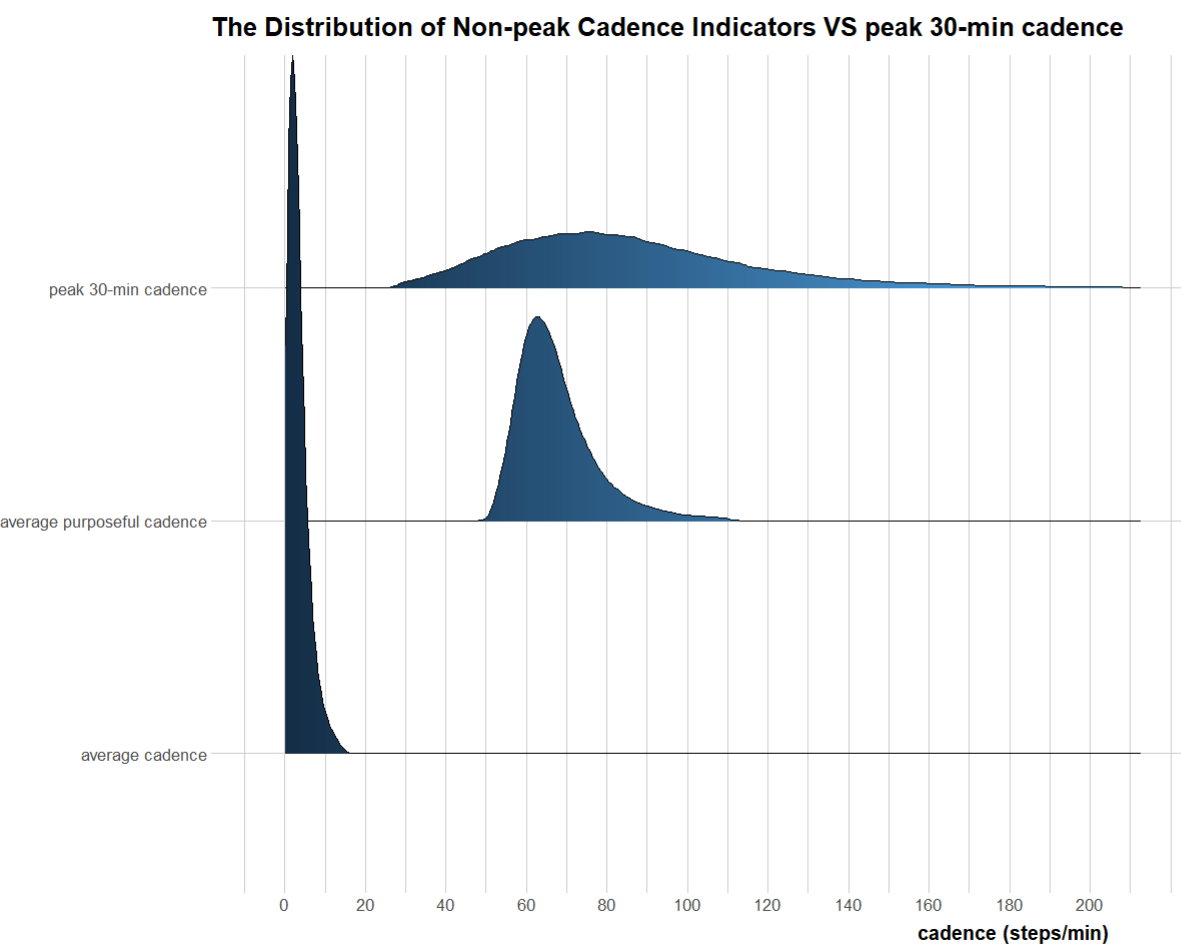

**b)**
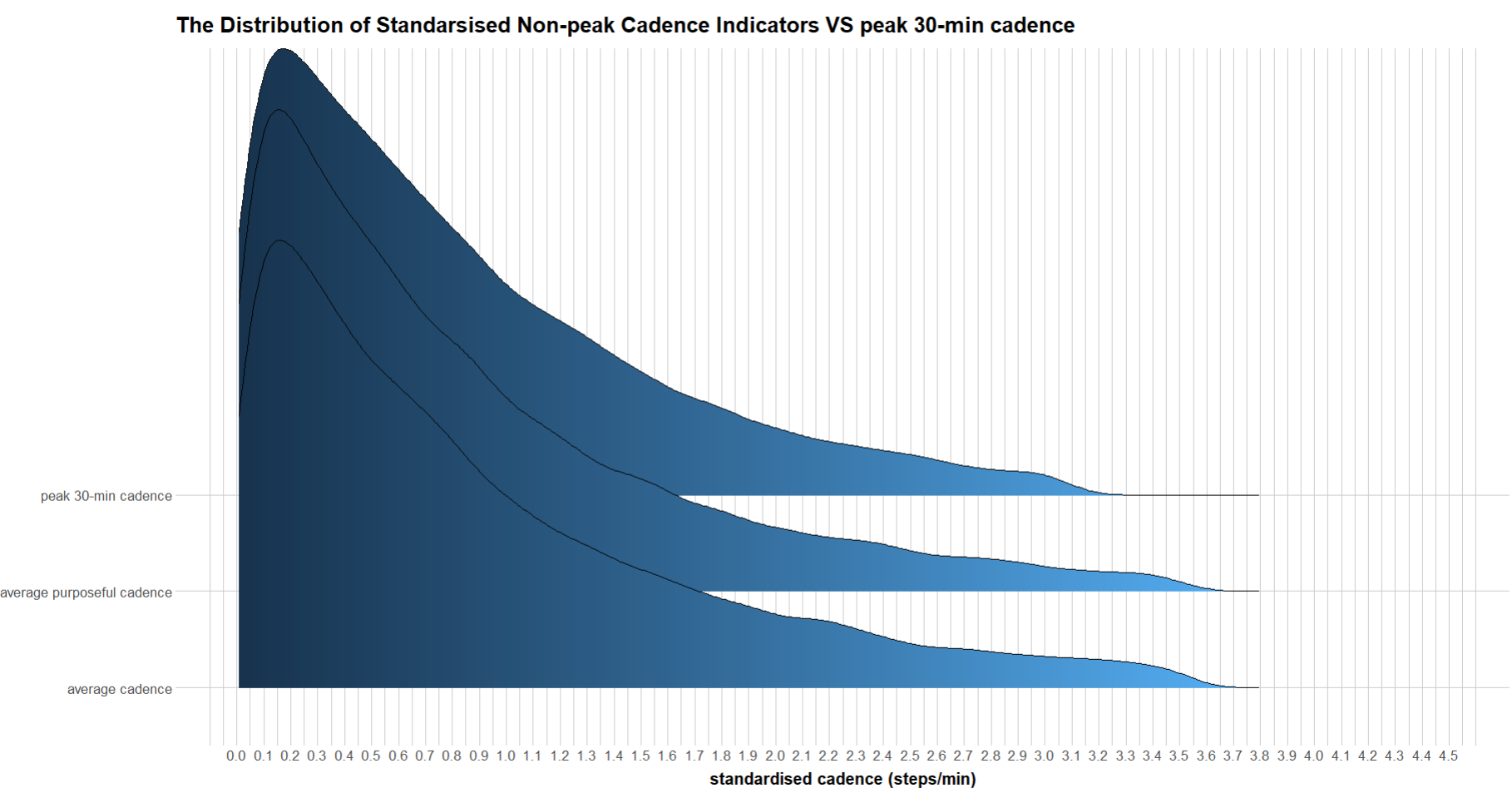

**Supplementary Fig. 4 Dose-Response Association of Standardised Stepping Intensity Estimated across Peak Cadence Metrics with Physical Activity-Related Cancer Mortality**

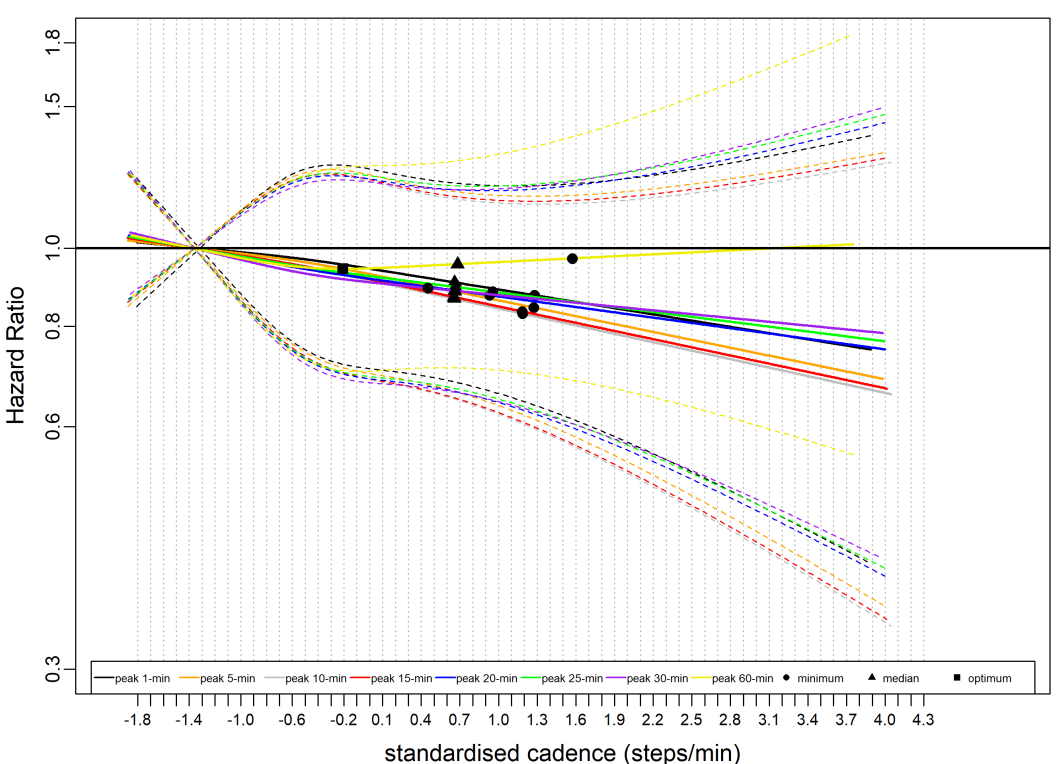

Total sample size is 70,174. The events for peak 1-min cadence is 406; peak 5-min cadence, 406; peak 10-min cadence, 407; peak 15-min cadence, 407; peak 20-min cadence, 408; peak 25-min cadence, 408; peak 30-min cadence, 409; peak 60-min cadence, 410. The circle indicates the ED50 value i.e., minimum, the minimal cadence associated with 50% of the optimal risk reduction; The triangle indicates the median cadence. The square indicates the cadence that associated with optimal mortality risk reduction (Note: the square was not annotated if there was no nadir point). We analysed the dose-response association using Fine and Grey model and adjusted for age, sex, accelerometer wearing duration, average daily steps, smoking status, alcohol consumption, sleep duration, Townsend deprivation score, sedentary time, education levels, self-reported parental history of CVD and cancer, and self-reported medication use (cholesterol, blood pressure, and diabetes). The reference level is 5^th^ percentile of the standardised distribution of each exposure.

**Supplementary Fig. 5 Dose-Response Association of Standardised Stepping Intensity Estimated by Non-peak Cadence Metrics (Average Cadence, Average Cadence of Purposeful Steps) and Peak 30-min Cadence with Physical Activity-Related Cancer Mortality**
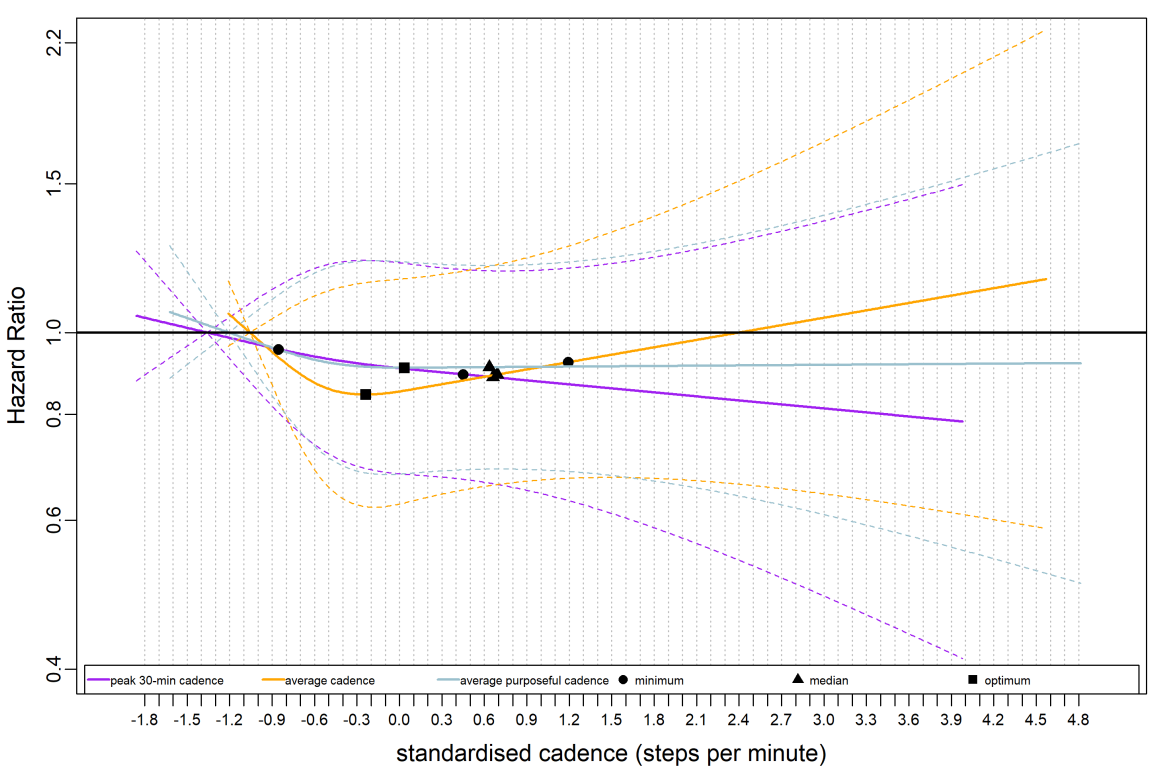

We compared two non-peak cadence metrics and a representative peak cadence metric (peak 30-min cadence) in this figure. The circle indicates the ED50 value i.e., minimum, the minimal standardised cadence that associated with 50% of the optimal risk reduction; The triangle indicates the median standardised cadence. The square indicates the standardised cadence that associated with optimal mortality risk reduction. We analysed the dose-response association using Fine and Grey model and adjusted for age, sex, accelerometer wearing duration, average daily steps, smoking status, alcohol consumption, sleep duration, Townsend deprivation score, sedentary time, education levels, self-reported parental history of CVD and cancer, and self-reported medication use (cholesterol, blood pressure, and diabetes). The reference level is 5^th^ percentile of the standardised distribution of each exposure.

**Supplementary Fig. 6 Dose-Response Association of Stepping Intensity Estimated across Peak Cadence Metrics with All-Cause Mortality**

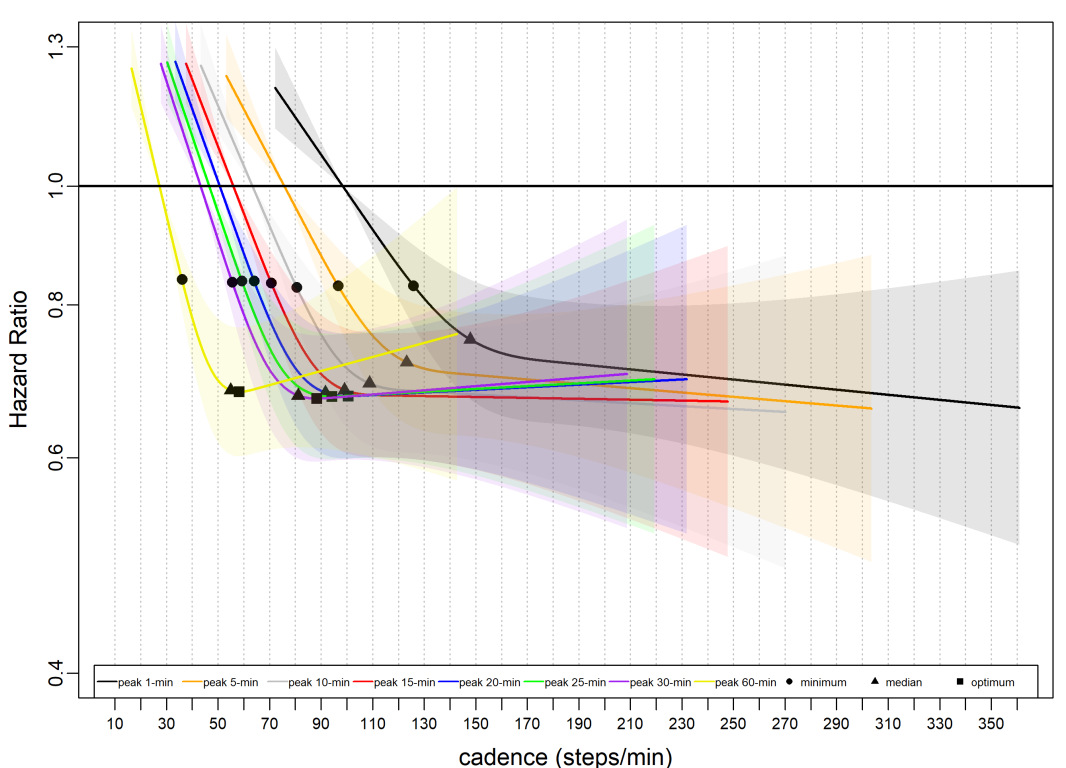

Total sample size is 70,174. The events for peak 1-min cadence is 2,029; peak 5-min cadence, 2,023; peak 10-min cadence, 2,019; peak 15-min cadence, 2,021; peak 20-min cadence, 2,022; peak 25-min cadence, 2,020; peak 30-min cadence, 2,021; peak 60-min cadence, 2,029. The circle indicates the ED50 value i.e., minimum, the cadence associated with 50% of the optimal risk reduction; The triangle indicates the median cadence. The square indicates the cadence that associated with optimal mortality risk reduction (Note: the square was not annotated if there was no nadir point). The dose-response association was analysed using cox-regression model and adjusted for age, sex, accelerometer wearing duration, average daily steps, smoking status, alcohol consumption, sleep duration, townsend deprivation score, sedentary time, education levels, self-reported parental history of CVD and cancer, and self-reported medication use (cholesterol, blood pressure, and diabetes). The reference level is 5^th^ percentile of each peak cadence metric.

**Supplementary Fig. 7 Dose-Response Association of Stepping Intensity Estimated by the Non-peak Cadence Metrics (Average Cadence, Average Cadence of Purposeful Steps) and Peak 30-min Cadence with All-Cause Mortality**

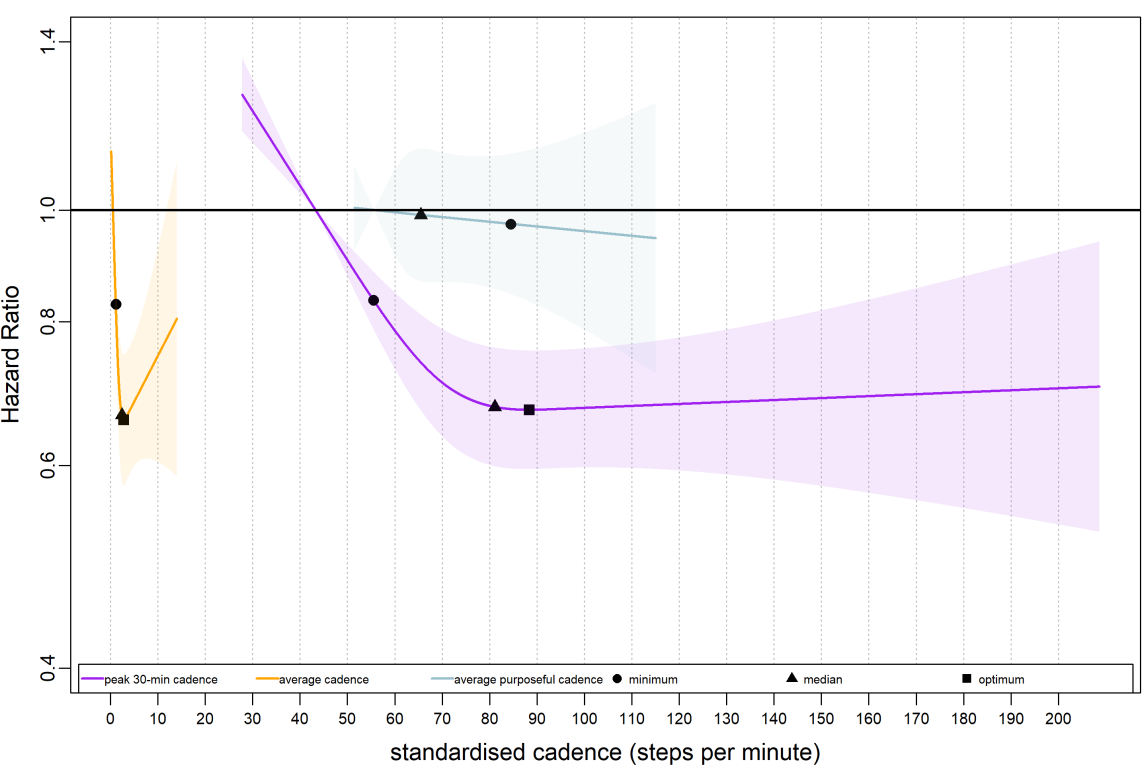

We compared two non-peak cadence metrics and a representative peak cadence metric (peak 30-min cadence) in this figure. Total sample size is 70,174. The events for peak 30-min cadence is 2,021; average cadence, 2,022; average cadence of purposeful steps, 2,064. The circle indicates the ED50 value i.e., minimum, the minimal steps per min associated with 50% of the optimal risk reduction; The triangle indicates the median cadence. The square indicates the cadence that associated with optimal mortality risk reduction (Note: the square was not annotated if there was no nadir point). The dose-response association was analysed using cox-regression model and adjusted for age, sex, accelerometer wearing duration, average daily steps, smoking status, alcohol consumption, sleep duration, Townsend deprivation score, sedentary time, education levels, self-reported parental history of CVD and cancer, and self-reported medication use (cholesterol, blood pressure, and diabetes). The reference level is 5^th^ percentile of the distribution of each exposure.

**Supplementary Fig. 8 Dose-Response Association of Stepping Intensity Estimated across Peak Cadence Metrics with CVD Mortality**

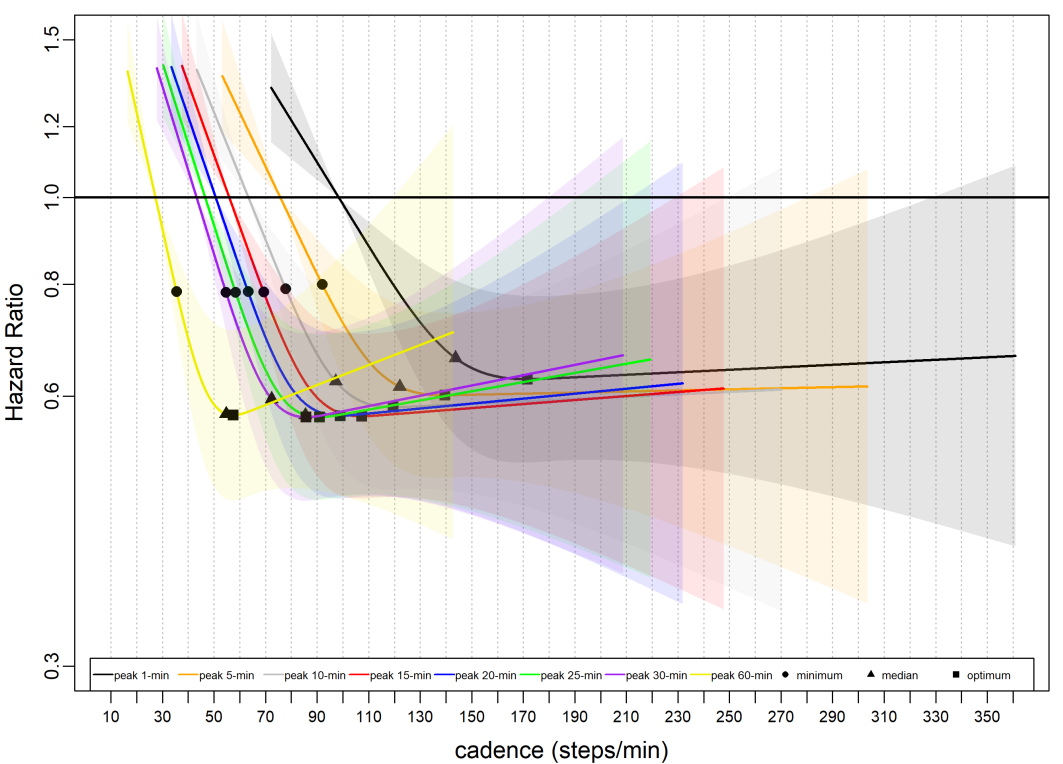

Total sample size is 70,174. The events for peak 1-min cadence is 549; peak 5-min cadence, 547; peak 10-min cadence, 546; peak 15-min cadence, 546; peak 20-min cadence, 546; peak 25-min cadence, 546; peak 30-min cadence, 548; peak 60-min cadence, 553. The triangle indicates the median cadence. The circle indicates the ED50 value i.e., minimum, the cadence associated with 50% of the optimal risk reduction; The square indicates the cadence that associated with optimal mortality risk reduction (Note: the square was not annotated if there was no nadir point). We used Fine and Grey model to analyse the dose-response association and adjusted for age, sex, accelerometer wearing duration, average daily steps, smoking status, alcohol consumption, sleep duration, townsend deprivation score, sedentary time, education levels, self-reported parental history of CVD and cancer, and self-reported medication use (cholesterol, blood pressure, and diabetes). The reference level is 5^th^ percentile of each peak cadence metric.

**Supplementary Fig. 9 Dose-Response Association of Stepping Intensity Estimated by Non-peak Cadence Metrics (Average Cadence, Average Cadence of Purposeful Steps) and Peak 30-min Cadence with CVD mortality**

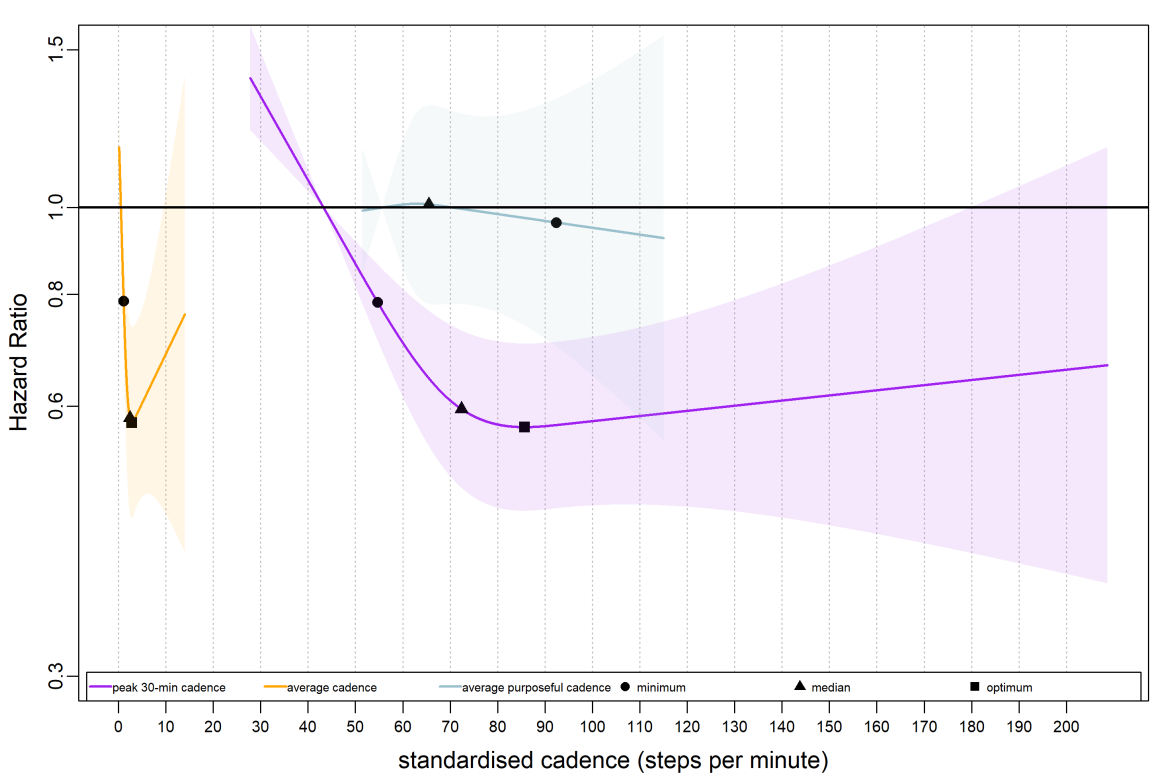

We compared two non-peak cadence metrics and a representative peak cadence metric (peak 30-min cadence) in this figure. Total sample size is 70,174, the events for peak 30-min cadence is 548; average cadence per day, 545; average cadence of purposeful steps per day, 569; purposeful cadence per day, 1,214. The circle indicates the ED50 value i.e., minimum, the minimal steps per min associated with 50% of the optimal risk reduction; The triangle indicates the median cadence. The square indicates the cadence that associated with optimal mortality risk reduction (Note: the square was not annotated if there was no nadir point). We used Fine and Grey model to analyse dose-response association and adjusted for age, sex, accelerometer wearing duration, average daily steps, smoking status, alcohol consumption, sleep duration, Townsend deprivation score, sedentary time, education levels, self-reported parental history of CVD and cancer, and self-reported medication use (cholesterol, blood pressure, and diabetes). The reference level is 5^th^ percentile of the distribution of each exposure.

**Supplementary** **Fig. 10 Dose-Response Association of Standardised Stepping Intensity Estimated across Peak Cadence Metrics with Cancer Mortality**

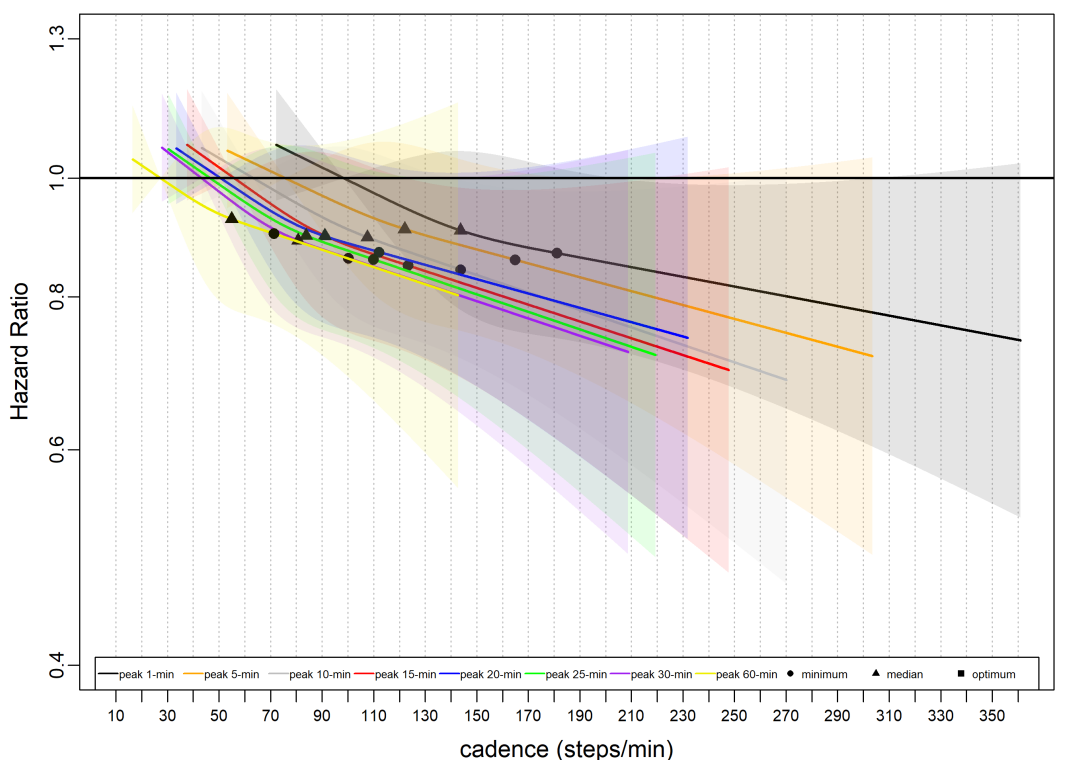

Total sample size is 70,174, the events for peak 1-min cadence is 1,205; peak 5-min cadence, 1,204; peak 10-min cadence, 1,201; peak 15-min cadence,1,201; peak 20-min cadence, 1,203; peak 25-min cadence, 1,201; peak 30-min cadence, 1,201; peak 60-min cadence, 1,201. The circle indicates the ED50 value i.e., minimum, the minimal steps per min associated with 50% of the optimal risk reduction; The triangle indicates the median steps per min. The square indicates the cadence that associated with optimal mortality risk reduction (Note: the square was not annotated if there was no nadir point). We used Fine and Grey model to analyse the dose-response association and adjusted for age, sex, accelerometer wearing duration, average daily steps, smoking status, alcohol consumption, sleep duration, townsend deprivation score, sedentary time, education levels, self-reported parental history of CVD and cancer, and self-reported medication use (cholesterol, blood pressure, and diabetes). The reference level is 5^th^ percentile of each peak cadence metric.

**Supplementary Fig. 11 Dose-Response Association of Stepping Intensity Estimated by Non-peak Cadence Metrics (Average Cadence, Average Cadence of Purposeful Steps) and Peak 30-min Cadence with Cancer Mortality**

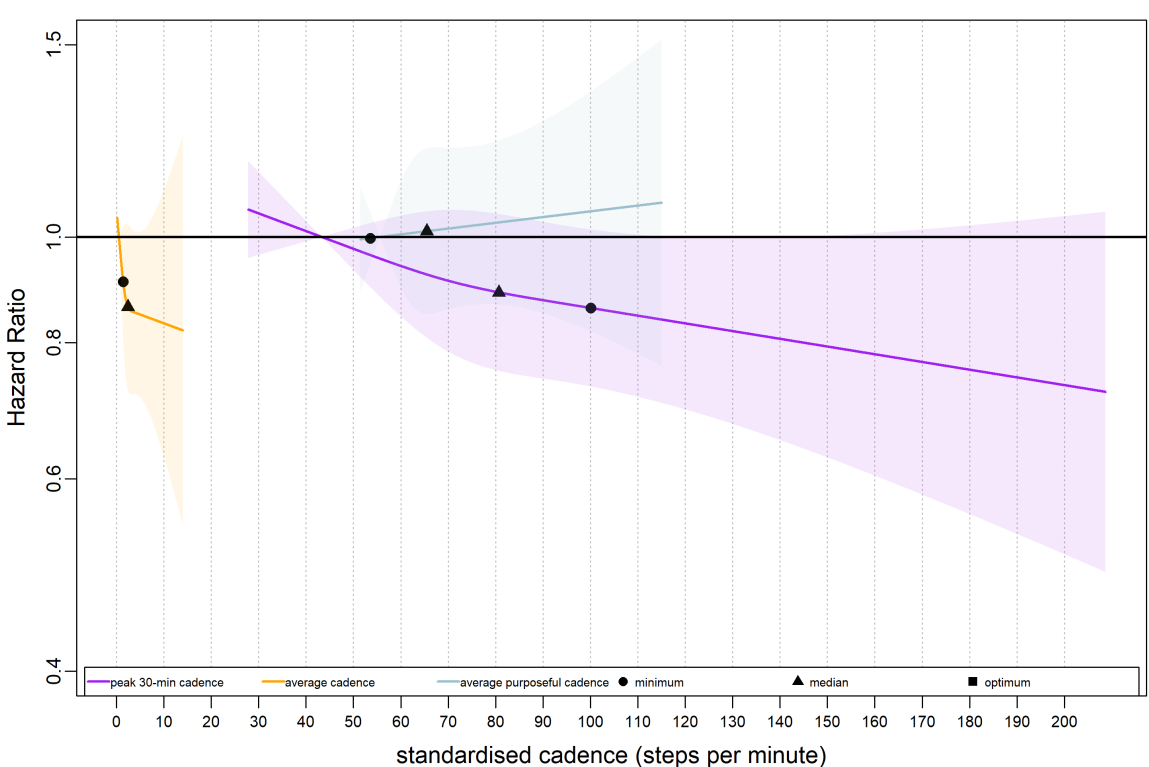

We compared two non-peak cadence metrics and a representative peak cadence metric (peak 30-min cadence) in this figure. Total sample size is 70,174. The events for peak 30-min cadence is 1,201; average cadence per day, 1205; average cadence of purposeful steps, 1214. The circle indicates the ED50 value i.e., minimum, the minimal cadence associated with 50% of the optimal risk reduction; The triangle indicates the median cadence. The square indicates the cadence that associated with optimal mortality risk reduction (Note: the square was not annotated if there was no nadir point). We analysed the dose-response association using Fine and Grey model and adjusted for age, sex, accelerometer wearing duration, average daily steps, smoking status, alcohol consumption, sleep duration, Townsend deprivation score, sedentary time, education levels, self-reported parental history of CVD and cancer, and self-reported medication use (cholesterol, blood pressure, and diabetes). The reference level is 5^th^ percentile of the distribution of each exposure.

**Supplementary Fig. 12 Dose-Response Association of Stepping Intensity Estimated across Peak Cadence Metrics with Physical Activity-Related Cancer Mortality**

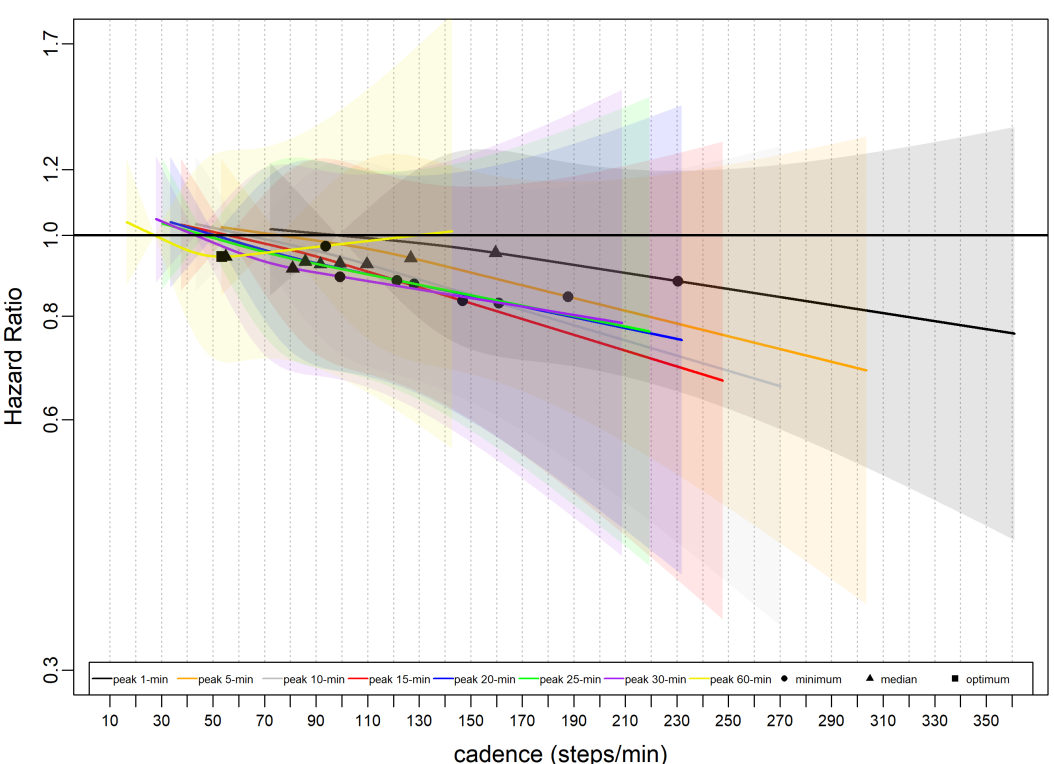

Total sample size is 70,174. The events for peak 1-min cadence is 406; peak 5-min cadence, 406; peak 10-min cadence, 407; peak 15-min cadence, 407; peak 20-min cadence, 408; peak 25-min cadence, 408; peak 30-min cadence, 409; peak 60-min cadence, 410. The circle indicates the ED50 value i.e., minimum, the minimal steps per min associated with 50% of the optimal risk reduction; The triangle indicates the median cadence. The square indicates the cadence that associated with optimal mortality risk reduction (Note: the square was not annotated if there was no nadir point). We analysed the dose-response association using Fine and Grey model and adjusted for age, sex, accelerometer wearing duration, average daily steps, smoking status, alcohol consumption, sleep duration, Townsend deprivation score, sedentary time, education levels, self-reported parental history of CVD and cancer, and self-reported medication use (cholesterol, blood pressure, and diabetes). The reference level is 5^th^ percentile of the standardised distribution of each exposure.

**Supplementary Fig. 13 Dose-Response Association of Stepping Intensity Estimated by Non-peak Cadence Metrics (Average Cadence, Average Cadence of Purposeful Steps) and Peak 30-min Cadence with Physical Activity-Related Cancer Mortality**

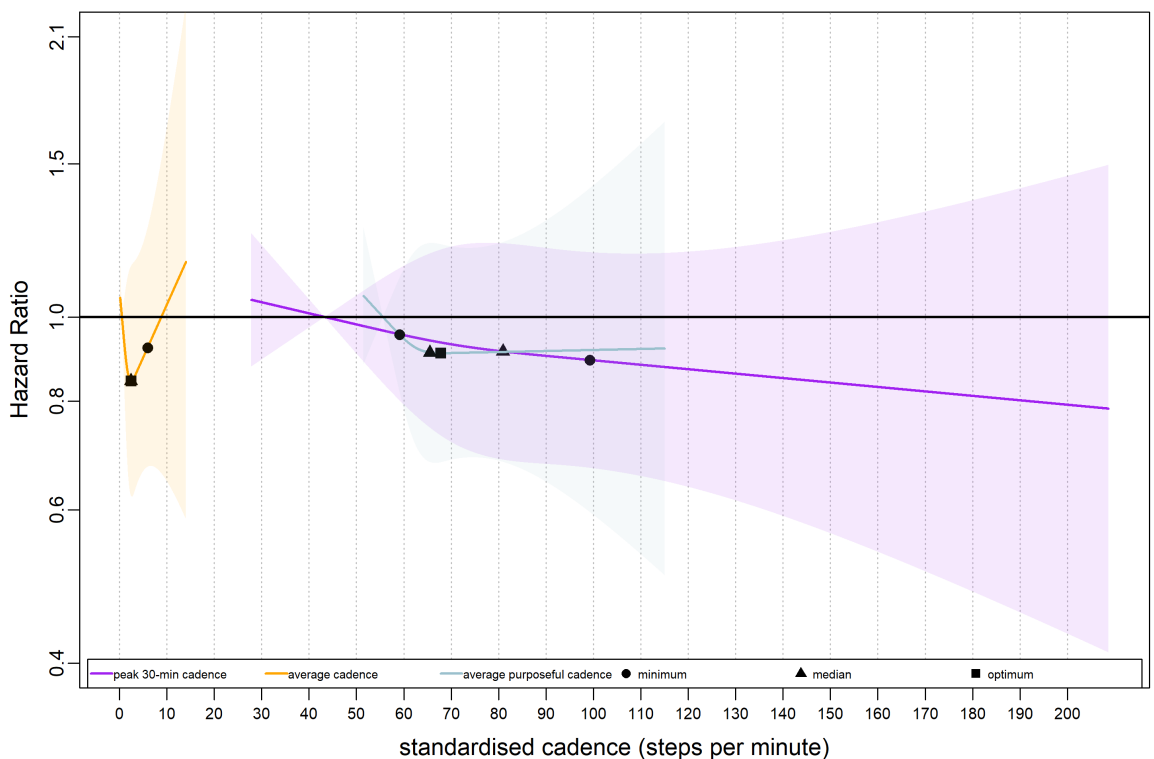

We compared two non-peak cadence metrics and a representative peak cadence metric (peak 30-min cadence) in this figure. The circle indicates the ED50 value i.e., minimum, the minimal cadence that associated with 50% of the optimal risk reduction; The triangle indicates the median cadence. The square indicates the cadence that associated with optimal mortality risk reduction. We analysed the dose-response association using Fine and Grey model and adjusted for age, sex, accelerometer wearing duration, average daily steps, smoking status, alcohol consumption, sleep duration, Townsend deprivation score, sedentary time, education levels, self-reported parental history of CVD and cancer, and self-reported medication use (cholesterol, blood pressure, and diabetes). The reference level is 5^th^ percentile of the standardised distribution of each exposure.

**Supplementary Fig. 14 Dose-Response Association of Standardised Stepping Intensity Estimated by Peak Cadence Metrics with All-cause Mortality Using Knots at 10^th^, 50^th^, and 90^th^ Percentile of Cadence Distribution**
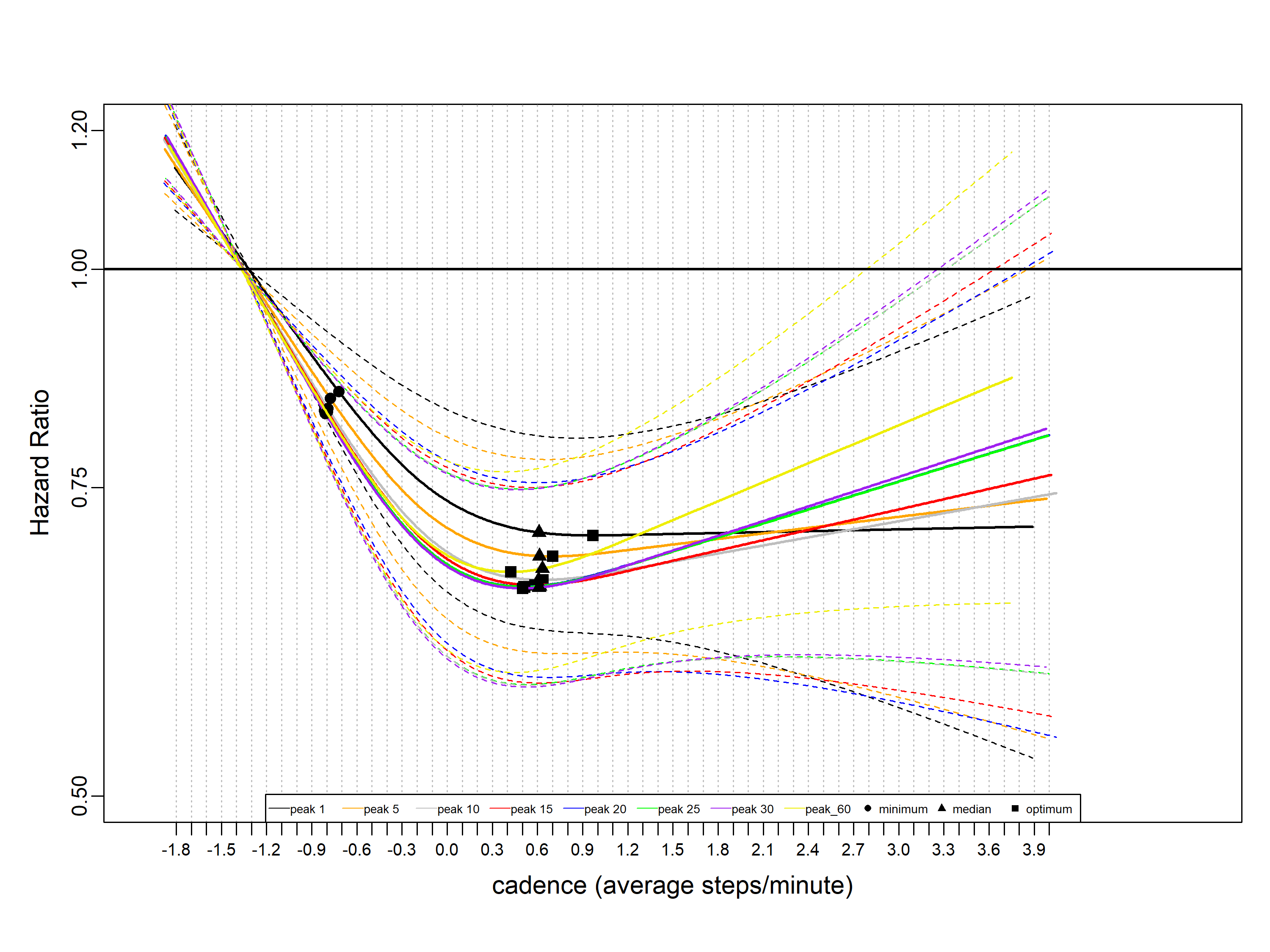

**Supplementary Fig. 15 Dose-Response Association of Normalised Stepping Intensity Estimated by Peak Cadence Metrics with All-cause Mortality**

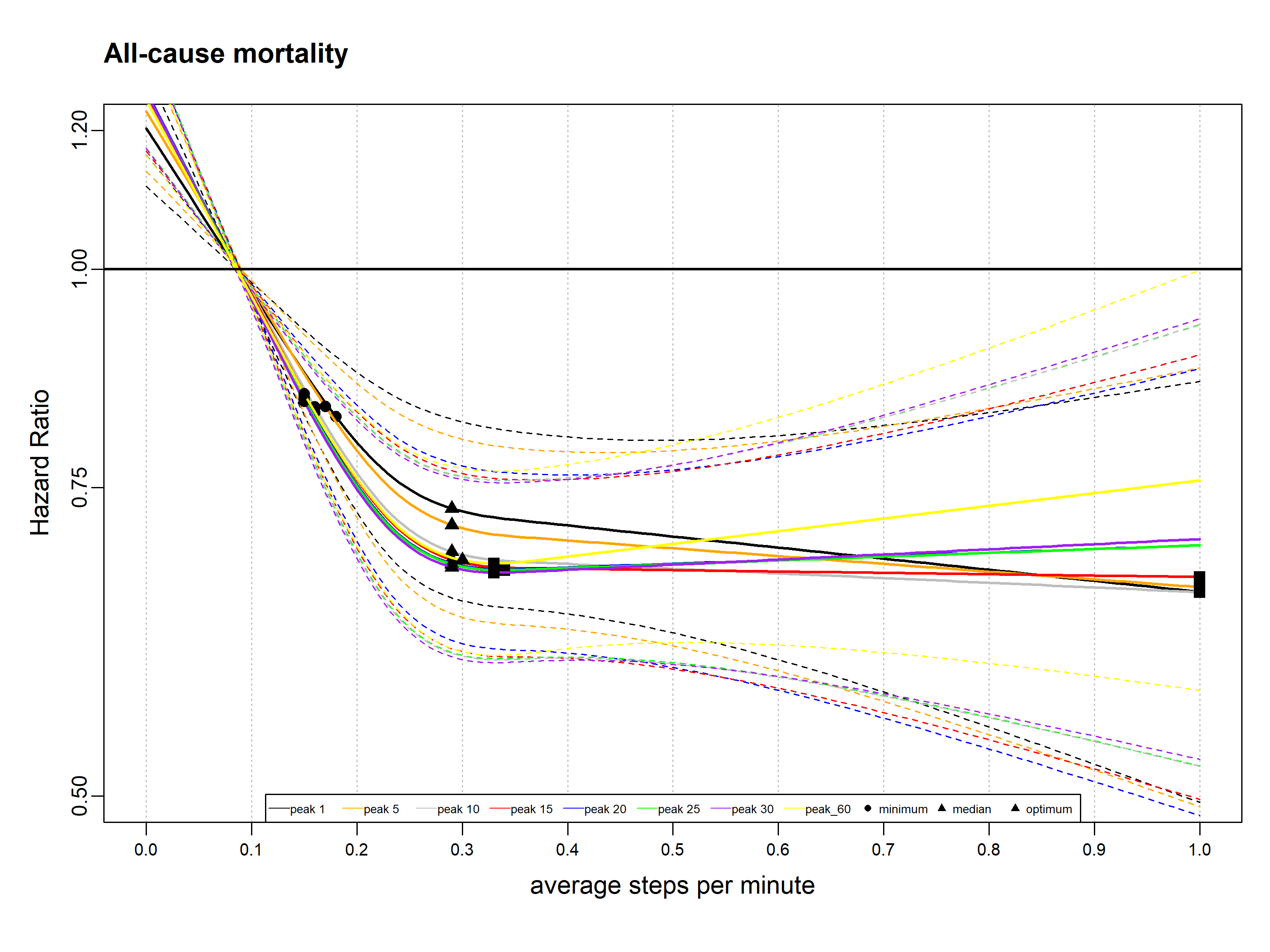

**Supplementary Fig. 16 Dose-Response Association of Standardised Stepping Intensity Estimated by Peak Cadence Metrics with All-cause Mortality after Removing the Outliers below 2.5^th^ and above 97.5^th^ Percentile**
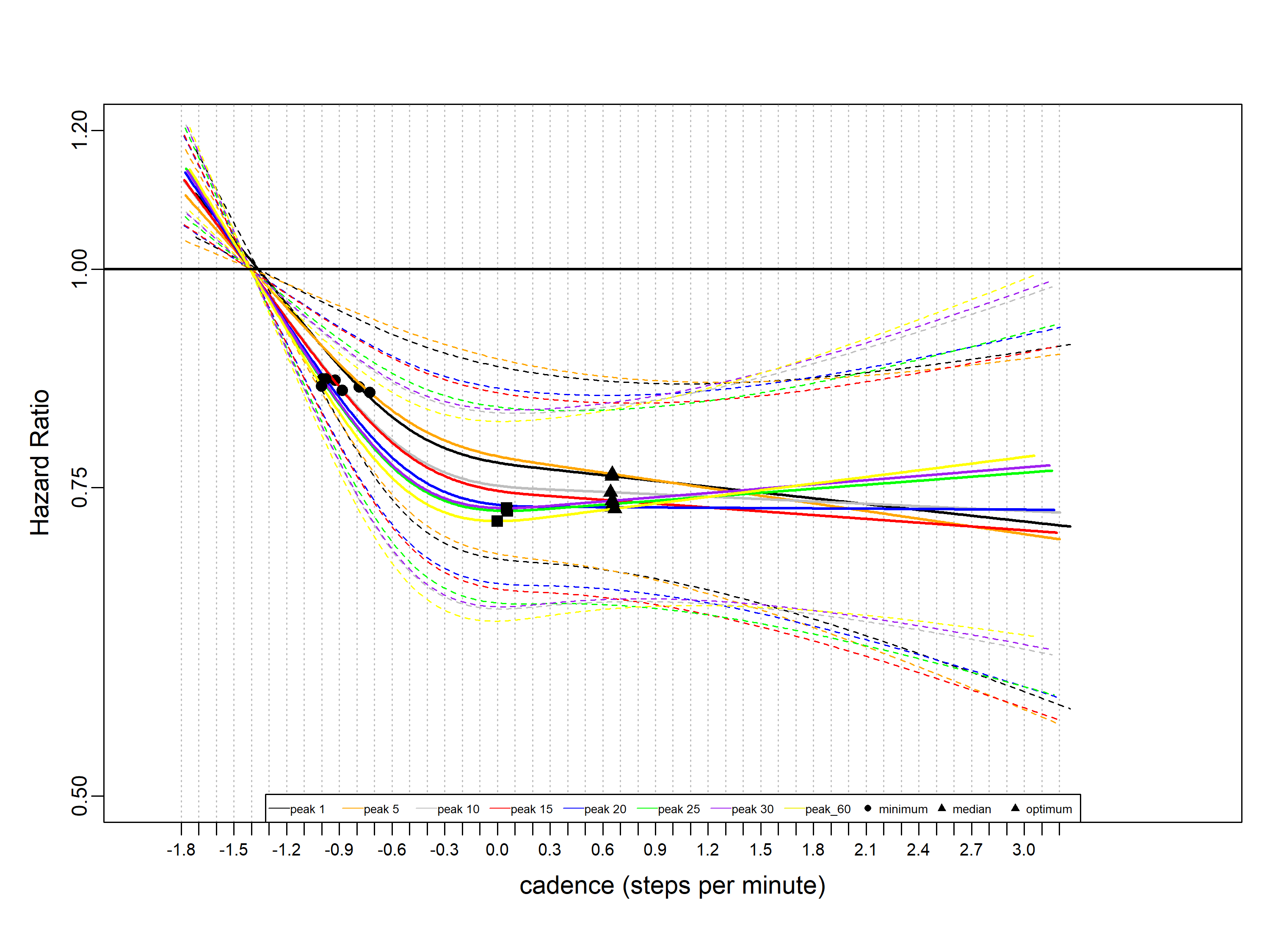

**Supplementary Table 1** Baseline Characteristics of Study Participants by Quartiles of Peak 30-min Cadence Per Day **^a^**

| **Characteristics** | **Overall** | **Bottom Quartile** | **Q2** | **Q3** | **Top Quartile** |
| --- | --- | --- | --- | --- | --- |
| Participants, No. | 70336 | 17584 | 17584 | 17584 | 17584 |
| Sex |  |  |  |  |  |
| Female | 40933 (58.2) | 11078 (63.0) | 10486 (59.6) | 10051 (57.2) | 9318 (53.0) |
| Male | 29403 (41.8) | 6506 (37.0) | 7098 (40.4) | 7533 (42.8) | 8266 (47.0) |
| Age | 61.6 (7.8) | 63.5 (7.6) | 62.00(7.7) | 61.0 (7.7) | 59.9 (7.8) |
| Accelerometer wearing days | 6.89 (0.39) | 6.9 (0.40) | 6.9 (0.4) | 6.9 (0.4) | 6.9 (0.4) |
| Follow-up years | 8.0 (0.9) | 7.9 (0.9) | 8.0 (0.9) | 8.0 (0.8) | 8.0 (0.8) |
| Stepping intensity metrics **^b^** |  |  |  |  |  |
| Peak 1-min cadence | 169.3 (55.9) | 33.3 (6.6) | 48.7 (4.9) | 62.4 (6.4) | 88.7 (17.9) |
| Peak 5-min cadence | 133.5 (43.3) | 51.0 (8.6) | 72.1 (5.1) | 90.8 (6.1) | 127.7 (22.8) |
| Peak 10-min cadence | 115.4 (38.5) | 54.7 (9.1) | 76.7 (5.3) | 96.0 (6.4) | 134.4 (23.9) |
| Peak 15-min cadence | 104.5 (35.8) | 59.2 (9.7) | 82.3 (5.9) | 102.4 (7.2) | 142.5 (25.4) |
| Peak 20-min cadence | 96.6 (33.8) | 65.2 (10.6) | 89.5 (7.0) | 110.6 (8.6) | 152.6 (27.5) |
| Peak 25-min cadence | 90.4 (32.2) | 73.7 (12.2) | 99.6 (9.0) | 121.9 (11.4) | 166.3 (30.8) |
| Peak 30-min cadence | 85.4 (30.9) | 88.4 (15.9) | 116.6 (14.1) | 140.6 (17.7) | 188.3 (37.1) |
| Peak 60-min cadence | 58.3 (22.9) | 118.9(29.0) | 151.1 (30.2) | 177.9 (36.1) | 229.2 (53.7) |
| Average_cadence | 3.1 (2.6) | 0.9 (0.4) | 1.9 (0.5) | 3.3 (1.0) | 6.3 (2.9) |
| Average cadence of purposeful_steps | 67.9 (11.8) | 67.9 (11.8) | 67.7(11.6) | 67.9 (11.7) | 68.0 (11.9) |
| Steps/day | 8135.2 (4635.5) | 3817.0 (1214.0) | 6335.7 (1583.8) | 8837.2 (2390.3) | 13551.0 (4954.6) |
| Townsend Deprivation Score **^c^** | -1.8 (2.8) | -1.7 (2.8) | -1.9 (2.8) | -1.8 (2.8) | -1.7 (2.8) |
| Alcohol_status, No. (%) **^d^** |  |  |  |  |  |
| Above_guideline | 26103 (37.1) | 5996 (34.1) | 6471 (36.8) | 6705 (38.1) | 6931 (39.4) |
| Never | 1933 (2.7) | 566 (3.2) | 481 (2.7) | 456 (2.6) | 430 (2.4) |
| Previous | 1744 (2.5) | 471 (2.7) | 434 (2.5) | 433 (2.5) | 406 (2.3) |
| Under_guideline | 40556 (57.7) | 10551 (60.0) | 10198 (58.0) | 9990 (56.8) | 9817 (55.8) |
| Fruit and vegetable servings/day | 8.0 (4.4) | 7.8 (4.4) | 7.9 (4.3) | 8.0 (4.3) | 8.2 (4.6) |
| Sleep hours/day | 7.5 (1.3) | 7.4 (1.5) | 7.5 (1.3) | 7.5 (1.2) | 7.5 (1.2) |
| Sedentary mins/day | 711.7 (93.4) | 761.5 (85.9) | 723.7 (79.0) | 696.3 (83.0) | 665.4 (96.9) |
| College or University degree, No. (%) | 31635 (45.0) | 7425 (42.2) | 7709 (43.8) | 7979 (45.4) | 8522 (48.5) |
| Cholesterol medication, No. (%) | 7686 (10.9) | 2701 (15.4) | 2030 (11.5) | 1660 (9.4) | 1295 (7.4) |
| Hypertension medication, No. (%) | 5693 (8.1) | 1836 (10.4) | 1518 (8.6) | 1264 (7.2) | 1075 (6.1) |
| Insulin medication, No. (%) | 78 (0.1) | 19 (0.1) | 21 (0.1) | 19 (0.1) | 19 (0.1) |
| Smoking_status, No. (%) |  |  |  |  |  |
| Never | 41319 (58.7) | 9915 (56.4) | 10242 (58.2) | 10479 (59.6) | 10683 (60.8) |
| Previous | 24464 (34.8) | 6331 (36.0) | 6172 (35.1) | 6028 (34.3) | 5933 (33.7) |
| Current | 4553 (6.5) | 1338 (7.6) | 1170 (6.7) | 1077 (6.1) | 968 (5.5) |
| Family history of CVD, No. (%) | 38206 (54.3) | 9967 (56.7) | 9608 (54.6) | 9465 (53.8) | 9166 (52.1) |
| Family history of cancer, No. (%) | 21725 (30.9) | 5490 (31.2) | 5584 (31.8) | 5364 (30.5) | 5287 (30.1) |
| All-cause mortality, No. (%) | 2037 (2.9) | 743 (4.2) | 513 (2.9) | 400 (2.3) | 381 (2.2) |
| CVD mortality, No. (%) | 553 (0.8) | 222 (1.3) | 130 (0.7) | 100 (0.6) | 101 (0.6) |
| Cancer mortality, No. (%) | 1209 (1.7) | 391 (2.2) | 339 (1.9) | 243 (1.4) | 236 (1.3) |
| PA_cancer mortality, No. (%) | 409 (0.6) | 128 (0.7) | 113 (0.6) | 79 (0.4) | 89 (0.5) |

Abbreviations: CVD, cardiovascular disease; PA, physical activity.

**^a.^** cadence was defined as steps per min.

**^b.^** Values represent mean (SD), unless specified otherwise.

**^c^** Lower score indicates more affluence

**^d^** Guidelines for alcohol use in the UK recommend no more than 14 units (1 unit = 10mL of pure alcohol) per week for both men and women.

**Supplementary Table 2** Hazard Ratio of All-Cause Mortality Associated with the Minimum, Median and Maximum Stepping Intensities (in Absolute Cadence Value), Estimated by Peak Cadence and Non-Peak Cadence Metrics

| **Stepping intensity** | minimum cadence | | median cadence | | maximum cadence | |
| --- | --- | --- | --- | --- | --- | --- |
|  | steps/min | HR (95% CI) | steps/min | HR (95% CI) | steps/min | HR (95% CI) |
| **Peak cadence metrics** |  |  |  |  |  |  |
| Peak 1-min cadence | 126.4 | 0.83 (0.77, 0.89) | 147.9 | 0.75 (0.67, 0.84) | 377.0 | 0.65 (0.50, 0.86) |
| Peak 5-min cadence | 96.6 | 0.83 (0.77, 0.89) | 123.2 | 0.72 (0.64, 0.81) | 303.6 | 0.66 (0.49, 0.88) |
| Peak 10-min cadence | 80.5 | 0.83 (0.78, 0.88) | 108.7 | 0.68 (0.61, 0.77) | 270.2 | 0.65 (0.49, 0.88) |
| Peak 15-min cadence | 70.7 | 0.83 (0.79, 0.88) | 99.1 | 0.68 (0.60, 0.77) | 247.7 | 0.67 (0.50, 0.89) |
| Peak 20-min cadence | 64.0 | 0.84 (0.79, 0.89) | 91.7 | 0.68 (0.60, 0.76) | 100.5 | 0.67 (0.60, 0.76) |
| Peak 25-min cadence | 59.3 | 0.84 (0.79, 0.89) | 86.7 | 0.67 (0.60, 0.75) | 94.2 | 0.67 (0.60, 0.75) |
| Peak 30-min cadence | 55.4 | 0.84 (0.79, 0.88) | 80.9 | 0.67 (0.60, 0.76) | 88.3 | 0.67 (0.60, 0.76) |
| Peak 60-min cadence | 36.0 | 0.84 (0.79, 0.89) | 54.9 | 0.68 (0.60, 0.77) | 58.3 | 0.68 (0.60, 0.77) |
| **Non-Peak cadence metrics** |  |  |  |  |  |  |
| Average cadence of purposeful steps | 86.0 | 0.96 (0.83, 1.11) | 65.5 | 0.99 (0.87, 1.14) | 110.5 | 0.92 (0.71, 1.19) |
| Average cadence | 1.15 | 0.83 (0.78, 0.88) | 2.42 | 0.67 (0.59, 0.76) | 2.84 | 0.66 (0.58, 0.76) |

**Supplementary Table 3** Hazard Ratio of CVD Mortality Associated with the Minimum, Median, and Maximum Stepping Intensities (in Absolute Cadence Value), Estimated by Peak Cadence and Non-Peak Cadence Metrics

| **Stepping intensity** | minimum cadence | | median cadence | | maximum cadence | |
| --- | --- | --- | --- | --- | --- | --- |
|  | steps/min | HR (95% CI) | steps/min | HR (95% CI) | steps/min | HR (95% CI) |
| **Peak cadence metrics** |  |  |  |  |  |  |
| Peak 1-min cadence | 120.4 | 0.80 (0.71, 0.88) | 143.7 | 0.66 (0.54, 0.81) | 171.6 | 0.63 (0.51, 0.78) |
| Peak 5-min cadence | 91.9 | 0.80 (0.72, 0.88) | 122.0 | 0.62 (0.50, 0.75) | 139.5 | 0.60 (0.49, 0.74) |
| Peak 10-min cadence | 77.8 | 0.79 (0.72, 0.87) | 97.2 | 0.62 (0.51, 0.76) | 119.6 | 0.58 (0.47, 0.72) |
| Peak 15-min cadence | 69.2 | 0.78 (0.71, 0.87) | 70.1 | 0.61 (0.50, 0.74) | 107.4 | 0.57 (0.46, 0.70) |
| Peak 20-min cadence | 63.2 | 0.78 (0.71, 0.86) | 61.4 | 0.61 (0.49, 0.73) | 98.9 | 0.57 (0.46, 0.70) |
| Peak 25-min cadence | 58.3 | 0.78 (0.71, 0.86) | 85.4 | 0.57 (0.46, 0.71) | 90.9 | 0.57 (0.46, 0.70) |
| Peak 30-min cadence | 54.6 | 0.78 (0.71, 0.86) | 72.4 | 0.60 (0.49, 0.73) | 85.6 | 0.57 (0.46, 0.70) |
| Peak 60-min cadence | 35.5 | 0.79 (0.71, 0.87) | 54.7 | 0.57 (0.46, 0.71) | 57.4 | 0.57 (0.46, 0.71) |
| **Non-Peak cadence metrics** |  |  |  |  |  |  |
| Average cadence of purposeful steps | 90.0 | 0.92 (0.67, 1.25) | 65.5 | 1.02 (0.79, 1.32) | 110.5 | 0.84 (0.51, 1.38) |
| Average cadence | 1.13 | 0.79 (0.70, 0.88) | 2.42 | 0.59 (0.46, 0.75) | 2.86 | 0.58 (0.45, 0.74 |

**Supplementary Table 4** Hazard Ratio of Cancer Mortality Associated with the Minimum, Median and Maximum Stepping Intensities (in Absolute Cadence Value), Estimated by Peak Cadence and Non-Peak Cadence Metrics

| **Stepping intensity** | minimum cadence | | median cadence | | maximum cadence | |
| --- | --- | --- | --- | --- | --- | --- |
|  | steps/min | HR (95% CI) | steps/min | HR (95% CI) | steps/min | HR (95% CI) |
| **Peak cadence metrics** |  |  |  |  |  |  |
| Peak 1-min cadence | 187.8 | 0.86 (0.73, 1.01) | 187.8 | 0.86 (0.74, 1.01) | 377.0 | 0.73 (0.51, 1.03) |
| Peak 5-min cadence | 164.8 | 0.86 (0.73, 1.01) | 122.0 | 0.91 (0.77, 1.06) | 303.6 | 0.71 (0.49, 1.04) |
| Peak 10-min cadence | 143.8 | 0.84 (0.71, 0.99) | 107.6 | 0.89 (0.76, 1.05) | 270.2 | 0.68 (0.46,1.00) |
| Peak 15-min cadence | 123.3 | 0.85 (0.72, 0.99) | 247.7 | 0.69 (0.48, 1.02) | 247.7 | 0.70 (0.48, 1.02) |
| Peak 20-min cadence | 112.1 | 0.87 (0.74, 1.02) | 91.0 | 0.90 (0.76, 1.05) | 231.8 | 0.74 (0.51, 1.07) |
| Peak 25-min cadence | 109.9 | 0.86 (0.72, 1.01) | 72.1 | 0.90 (0.74, 1.07) | 219.5 | (0.72, 0.49, 1.04) |
| Peak 30-min cadence | 100.3 | 0.86 (0.72, 1.01) | 80.6 | 0.89 (0.75, 1.04) | 208.7 | 0.72 (0.49, 1.05) |
| Peak 60-min cadence | 71.2 | 0.90 (0.76, 1.06) | 53.3 | 0.88 (0.72, 1.07) | 142.8 | 0.80 (0.56, 1.15) |
| **Non-Peak cadence metrics** |  |  |  |  |  |  |
| Average cadence of purposeful steps | 53.6 | 0.99 (0.94, 1.05) | 65.5 | 1.01 (0.85, 1.21) | 51.5 | 0.99 (0.89, 1.10) |
| Average cadence | 1.23 | 0.92 (0.84, 1.01) | 2.42 | 0.86 (0.73, 1.02) | 14.0 | 0.85 (0.56, 1.29) |
